# A Bibliometric Review of Explainable AI in Diabetes Risk Prediction: Trends, Gaps, and Knowledge Graph Opportunities

**DOI:** 10.64898/2026.04.16.26351069

**Authors:** Thieu Anh Van

## Abstract

**Background:** Type 2 diabetes mellitus (T2DM) is a leading global public health challenge. Machine learning (ML) combined with Explainable AI (XAI) is increasingly applied to T2DM risk prediction, but the field lacks a quantitative overview of methodological trends and integration gaps.

**Methods:** We present a structured synthesis and critical analysis of the XAI literature on T2DM risk prediction, combining (i) quantitative bibliometric analysis of a two-database corpus (N = 2,048 documents from Scopus and PubMed/MEDLINE, deduplicated via a transparent three-tier pipeline) and (ii) an in-depth selective review of 15 highly cited papers. Reporting follows PRISMA 2020, adapted for metadata-based synthesis; analyses include keyword frequency, rule-based thematic clustering, and publication trend analysis.

**Results:** The field grew rapidly, from 36 documents (2020) to 866 (2025). SHAP and LIME dominate XAI methods; XGBoost and Random Forest dominate ML models. Critically, KG/GNN terms appeared in only 17 documents (∼0.83%) compared with 906 for XAI methods, a 53.3:1 disparity. This gap is consistent across both databases, which share 33.2% of their records, ruling out a single-database artifact. The selective review confirmed that none of the 15 highly cited papers combined all three components, ML, XAI, and KG, in T2DM risk prediction.

**Conclusions:** The XAI for T2DM risk prediction field exhibits a clinical interpretability gap: statistical explanations are rarely linked to structured clinical pathways. We propose a three-layer conceptual framework (Predictive → Explainability → Knowledge) that integrates KG as a supplementary semantic layer, with potential applications in clinical decision support and population-level screening. The framework does not perform true causal inference but structures explanations around established pathophysiological knowledge. This study contributes a transferable methodology and a quantified research gap to guide future work integrating ML, XAI, and structured medical knowledge.

## 1. Introduction

### 1.1 Research Motivation

Type 2 diabetes mellitus (T2DM) is one of the most serious public health challenges of the 21st century. According to the International Diabetes Federation (IDF), T2DM has reached alarming global proportions, with particularly rapid growth in low- and middle-income countries; 589 million adults currently live with the disease, equivalent to 11.1% of the global adult population, and this figure is projected to rise to 852.5 million by 2050 (3). The economic burden exceeded USD 1 trillion in healthcare costs in 2024, while diabetes caused more than 3.4 million deaths in the same year (3). Against this backdrop, early detection and population-level risk prediction represent urgent public health priorities.

The development of machine learning (ML) has opened the door to building T2DM risk prediction models. Large-scale population health datasets such as the Behavioral Risk Factor Surveillance System (BRFSS) have been widely used in prior studies to evaluate predictive models and XAI methods for T2DM (4). However, most complex ML models — especially XGBoost, Random Forest, and deep neural networks — operate as “black boxes”, generating predictions without transparent explanations. This creates a significant barrier to clinical deployment, where clinicians need not only predictive outcomes but also an understanding of why a patient is classified as high or low risk.

Explainable Artificial Intelligence (XAI) has been proposed as a solution to this limitation. Methods such as SHAP (5) and LIME (6) enable model predictions to be explained at the feature level, providing information about the contribution of each risk factor (BMI, blood pressure, lifestyle…) to the prediction outcome.

### 1.2 Problem Statement and Research Gaps

Although reviews exist on XAI in biomedicine (11), ML in T2DM (30), and KG in healthcare (8), each domain has its own rich literature — yet, to our knowledge, no study has quantified the gap at the intersection of all three. Specifically, within the T2DM research corpus, XAI methods show 906 keyword occurrences across 2,048 documents (Scopus + PubMed), while KG/GNN has only 17 (∼0.83%) — a 53.3:1 disparity. More importantly, within the corpus analyzed, no study simultaneously combines ML prediction, systematic XAI, and a KG semantic layer in a T2DM risk-prediction pipeline from population health data. This study fills that gap by quantifying it and proposing a structured integration framework.

### 1.3 Research Questions

Four research questions are operationalized as follows — each directly linked to a specific analytical step:

- RQ1 — Temporal trends: What is the publication growth rate for XAI in T2DM over 2015–2026, and are there signs of saturation? Measured by: publication year distribution.
- RQ2 — Geographic and source distribution: Which countries and journals lead? Measured by: affiliation and source title statistics.
- RQ3 — Prevalent methods: Which XAI methods and ML models are most common? Measured by: author keyword frequency by group.
- RQ4 — KG/GNN gap: What is the prevalence of KG/GNN in the corpus, and what are the practical implications? Measured by: KG/GNN vs. XAI keyword frequency, combined with a selective review of 15 papers.

### 1.4 Principal Contributions

This paper makes four principal contributions:

- (a) Provides a systematic bibliometric analysis of XAI in T2DM risk prediction on a two-database corpus of 2,048 documents (Scopus + PubMed/MEDLINE). The multi-database design addresses the validation requirement for bibliometric studies.
- (b) Identifies and validates the KG/GNN gap through quantitative analysis (0.83% in the two-database corpus) and selective review of 15 highly cited papers — a 53.3:1 disparity relative to XAI methods, strengthened by consistency across both databases (33.2% record overlap).
- (c) Proposes a three-layer conceptual framework (Predictive → Explainability → Knowledge) that integrates KG with XAI to supplement feature-level statistical explanations with structured pathophysiological pathway reasoning aligned with clinical ontologies (e.g., UMLS, ICD-10; illustrated in Fig. 1).
- (d) Identifies research gaps in clinically interpretable decision support systems based on structured clinical pathways — rather than purely statistical correlations — as a largely unexplored direction in this field.

**Fig. 1.**
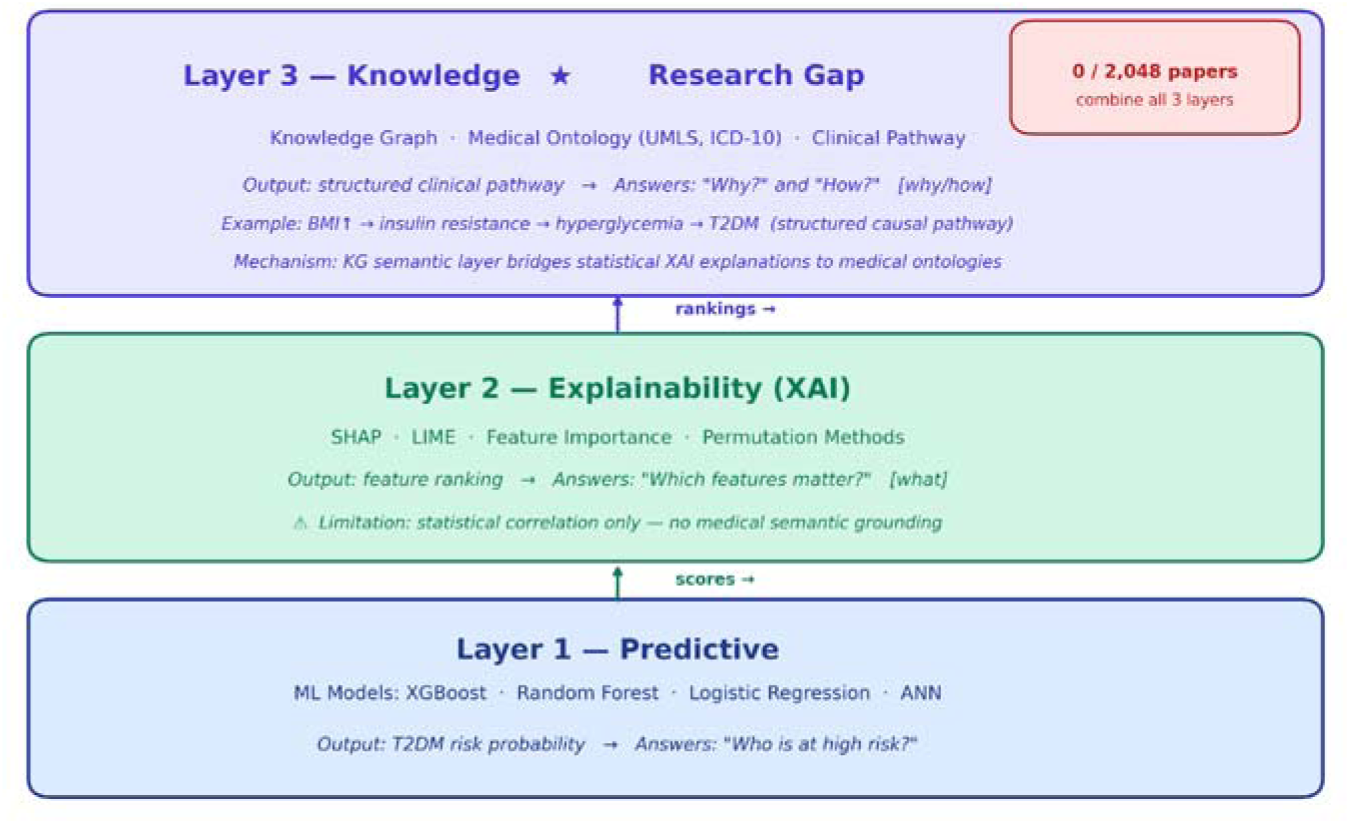
Three-layer conceptual framework proposed for integrating ML + XAI + KG in T2DM risk prediction.

Together, these four contributions position the study as a transferable methodology and a quantified empirical gap, setting the stage for the positioning against existing reviews in §1.5.

### 1.5 Positioning Relative to Existing Reviews

To clarify the unique contribution of this study, Table 1 compares this work with six representative reviews in related fields. Comparison criteria include: topic scope, method (quantitative bibliometric or narrative review), presence/absence of KG/GNN analysis, presence/absence of an integration framework, and corpus size.

**Table 1.** Comparison of this study with representative existing reviews. ✓ = present; ✗ = absent; ★ = current study. Corpus = number of documents analyzed.

| Study | Scope | Method | KG/GNN Analysis | Framework | Corpus |
| --- | --- | --- | --- | --- | --- |
| Tjoa & Guan (11) | XAI general/medical | Narrative review | No | No | N/A |
| Kavakiotis et al. (30) | ML in T2DM | Narrative review | No | No | N/A |
| Budhdeo et al. (8) | KG in biomedical | Scoping review | Yes (non-T2DM) | No | 255 |
| Kiran et al. (31) | ML/AI in T2DM (33 yr) | Bibliometric | No | No | Not stated |
| Kim et al. (33) | XAI-based CDSS | Systematic review | No | No | N/A |
| □ This study | XAI × T2DM risk | Bibliometric (quantitative) | Yes (0.83% gap quantified) | Yes (three-layer) | 2,048 (Scopus+PubMed) |

To the best of our knowledge, few prior studies have combined all of the following elements in a single analysis: (1) quantitative bibliometric analysis on a large two-database corpus (2,048 papers from Scopus and PubMed/MEDLINE); (2) a specific focus on XAI for T2DM risk prediction from population health data; (3) quantification of the KG/GNN gap (0.83%) rather than a purely qualitative mention; and (4) a proposed three-layer conceptual framework integrating ML+XAI+KG. Notably, Kiran et al. (31) also conducted a 33-year bibliometric analysis of ML and AI in T2DM prediction in the same journal Frontiers in Digital Health (2025) — however, that study does not separately analyze the XAI field, does not quantify the KG/GNN gap, and does not propose an integration framework, which we view as a complementary rather than overlapping contribution.

## 2. Background

### 2.1 Diabetes and Global Burden

Diabetes mellitus is a chronic metabolic disorder characterized by persistent hyperglycemia, arising from deficient insulin secretion, reduced insulin sensitivity, or both (9). Type 2 diabetes (T2DM) accounts for over 90% of total cases and is associated with modifiable risk factors, including overweight, obesity, physical inactivity, and unhealthy diet (3). According to IDF 2025, the epidemic will grow by 45% by 2050, with 95% of the increase concentrated in low- and middle-income countries — where health systems are most constrained (3). Serious complications include cardiovascular disease, chronic kidney disease, blindness, and limb amputation, placing a heavy burden on quality of life and global health systems (9).

### 2.2 Machine Learning in T2DM Risk Prediction

Machine learning has been widely applied to T2DM risk prediction. Ensemble methods — particularly Random Forest and XGBoost (1) — demonstrate superior performance on structured health data compared to traditional methods. XGBoost is a highly scalable gradient-boosting system with built-in regularization and efficient handling of sparse data (1). Deep learning has also been explored — for example, ensemble deep learning frameworks applied to multi-source diabetes data (46) — though its advantage over ensemble methods has been inconsistent on structured clinical data. Despite high accuracy, most models operate as black boxes — creating barriers to real-world clinical deployment.

### 2.3 XAI Taxonomy for T2DM Prediction

A taxonomy of XAI methods in T2DM prediction can be structured along three principal criteria (10–11):

#### By explanation scope

- Global explanation: explains the overall model behavior across the entire dataset — e.g., Feature Importance from Random Forest, SHAP summary plot.
- Local explanation: explains individual predictions for a specific patient — e.g., SHAP waterfall plot, LIME per-instance explanation.

#### By model dependency

- Model-specific: designed for a specific architecture — e.g., SHAP TreeExplainer for XGBoost/Random Forest.
- Model -agnostic: applicable to any model — e.g., LIME, Permutation Feature Importance, Kernel SHAP.

#### By sensitivity to feature correlation

- Correlation -aware: SHAP handles highly correlated features better, allocating contributions based on marginal contributions (Shapley values).
- Correlation-sensitive: Permutation Feature Importance and LIME are susceptible to noise when features are highly correlated.

XAI addresses the transparency limitations of ML models. SHAP (SHapley Additive exPlanations) (5), grounded in cooperative game theory, assigns each feature a marginal contribution value satisfying three properties: local accuracy, missingness, and consistency

— ensuring explanations are not contradicted by model logic. A specialized variant, TreeSHAP (38), computes explanations for tree-based models in polynomial time, making SHAP computationally feasible on large tabular clinical datasets. LIME (6) builds a simple linear model to locally approximate the behavior of a complex model around a specific data point. Hoyos et al. (7) demonstrated the value of combining SHAP, feature importance, and scenario simulation to produce a comprehensive T2DM analysis.

### 2.4 Current XAI Challenges in T2DM Prediction

Despite significant advances, XAI in T2DM faces the following fundamental challenges (44, 47):

- Challenge 1 — Correlated features distort explanations: when clinical features (BMI, blood pressure, cholesterol) are highly correlated, XAI methods such as Feature Importance may misallocate importance, leading to inconsistent and unreliable explanations.
- Challenge 2 — Lack of ground truth for XAI evaluation: there is no objective standard to evaluate whether an XAI explanation is clinically correct, unlike prediction accuracy, where ground truth labels exist. Current evaluations rely primarily on inter-method consistency rather than independent medical evidence.
- Challenge 3 — Clinical interpretability gap (12): statistical explanations from SHAP (“BMI contributes 0.32 points”) do not naturally align with how clinicians reason through pathological chains. This gap limits the practical applicability of XAI in clinical settings.
- Challenge 4 — Lack of cross-method agreement: different XAI methods (SHAP, LIME, Feature Importance) often produce inconsistent feature rankings on the same dataset. This complicates clinical decision-making — it is unclear which method to trust (11–12).

### 2.5 Limitations of XAI and the Role of Knowledge Graphs

Although SHAP and LIME substantially improve the transparency of ML models (5, 6, 10), they share a fundamental limitation: their explanations stop at the level of statistical feature attribution and cannot represent medical knowledge about the causal relationships among risk factors. SHAP can indicate that “BMI contributes 0.32 points to the predicted probability of diabetes,” but it cannot articulate why BMI is dangerous, nor trace the pathophysiological chain: obesity → insulin resistance → hyperglycemia → T2DM. In other words, current XAI answers the “what” question (which features matter) but not the “why” and “how” questions in a clinically meaningful sense.

This is the gap that knowledge graphs (KG) can help address. A KG represents medical knowledge as a directed graph of entities and structured semantic relationships (40) — for example, Lobesity, increases-risk-of, insulin resistanceL or LT2DM, causes-complication, cardiovascular diseaseL. In clinical contexts, graph-based patient representations have also been shown to support the integration of heterogeneous data (42). This structure lets a system move beyond prediction toward reasoning along logical chains grounded in structured causal knowledge, which aligns with the requirements of clinical decision support. Budhdeo et al. (8), in a scoping review of 255 studies, confirmed that KGs are increasingly applied in biomedicine. Do and Pham (14) proposed W-KG2Vec — a KG representation-learning model combining graph structure and textual semantic information — demonstrating the potential of KG embeddings for medical knowledge-related tasks (15). This approach also addresses a concern raised in clinician surveys (39), which report that feature-level explanations alone are often insufficient for clinical reasoning and that clinicians prefer explanations contextualized by structured clinical knowledge.

## 3. Methods

### 3.1 Data Source and Search Strategy

This study uses two independent scientific databases: Scopus and PubMed/MEDLINE. These two databases were selected as the most representative sources for this study’s scope: Scopus provides broad coverage (>100 million records) across computer science and engineering — where most XAI methodology research is published — while PubMed/MEDLINE is the standard biomedical and clinical database, essential given the diabetes subject matter. Together, they cover the intersection of computational methods (Scopus) and clinical/biomedical literature (PubMed) relevant to XAI in T2DM risk prediction, and satisfy the multi-database validation requirement for bibliometric studies. For both databases, the search strategy was constructed using two keyword groups combined with AND, and an exclusion group (NOT) to ensure corpus specificity:

Group 1 — domain (diabetes): “diabetes” OR “diabetes mellitus” OR “type 2 diabetes” OR “T2DM” OR “diabetic” OR “diabetes prediction” OR “diabetes risk”

Group 2 — method (XAI): “explainable AI” OR “XAI” OR “explainability” OR “interpretable machine learning” OR “interpretable AI” OR “SHAP” OR “LIME” OR “feature importance” OR “model interpretability” OR “explainable machine learning” OR “black-box model” OR “model transparency” OR “interpretable model”

Exclusion group (NOT): “retinopathy” OR “diabetic retinopathy” OR “fundus” OR “optical coherence” OR “OCT” OR “retinal” OR “macular” OR “optic disc” OR “diabetic foot” OR “wound healing” OR “skin lesion.”

Scopus search was executed in the TITLE-ABS-KEY field on 29 March 2026. PubMed search was executed in the [tiab] field (title/abstract, semantically equivalent to TITLE-ABS-KEY) via NCBI E-utilities API on 20 April 2026. The MeSH field [mh] was deliberately not used in PubMed to preserve semantic equivalence across the two queries. 2026 figures are therefore incomplete as the year has not ended. The exclusion group was constructed to remove two specialized sub-fields not relevant to XAI for risk prediction from structured clinical data: (1) diabetic eye complications (retinopathy, fundus, OCT…) — which use computer vision and medical image analysis, entirely different methodology from XAI for risk prediction from structured clinical data; and (2) diabetic foot and wound complications — focused on treatment and wound care, unrelated to T2DM onset risk prediction.

### 3.2 Inclusion and Exclusion Criteria

#### Inclusion criteria

- Documents indexed in Scopus or PubMed/MEDLINE without year restriction
- Language: unrestricted (predominantly English)
- Document types: research articles, conference papers, reviews, book chapters

#### Exclusion criteria

- Studies on diabetic eye complications (retinopathy, fundus, OCT, retinal, macular, optic disc) — excluded because this is a specialized sub-field using medical image analysis methodology (computer vision/deep learning for image classification), entirely distinct in methodology from XAI for risk prediction from structured clinical data
- Studies on diabetic wounds and skin lesions (diabetic foot, wound healing, skin lesions)
- Retracted documents, editorials, errata

### 3.3 Data Processing and Analysis Procedure

Records were retrieved from Scopus (CSV export with full metadata fields, 29 March 2026) and PubMed/MEDLINE (via NCBI E-utilities API (34) using Biopython (35), 20 April 2026). Both raw exports were harmonized to a common schema (title, authors, year, source, DOI, PMID, abstract, keywords, affiliations) before the three-tier deduplication described in Section 3.1 and illustrated in Fig. 2. Deduplicated data were processed through three main steps:

1. Data cleaning and normalization: (a) synonym merging: “explainability”, “explainable AI”, “XAI” → same group; “T2DM”, “type 2 diabetes”, “diabetes mellitus type 2” → same group; (b) normalize compound terms: “machine-learning” → “machine learning”; (c) convert to lowercase; remove non-English keywords from frequency analysis (retained for geographic analysis); (d) self-citations were not removed individually due to Scopus export limitations. The impact of self-citations is noted as a study limitation; however, the self-citation rate in bibliometric studies typically ranges 10–20%, so the effect on overall trend analysis is estimated to be negligible.
2. Quantitative analysis using Python (pandas 1.5+, matplotlib 3.5+, numpy): year distribution, geography, publication sources, keyword frequency, rule-based clustering, top-cited.
3. Selective review of 15 highly cited papers to qualitatively validate gaps identified from quantitative analysis. All code is publicly available (see Data Availability Statement).

**Fig. 2.**
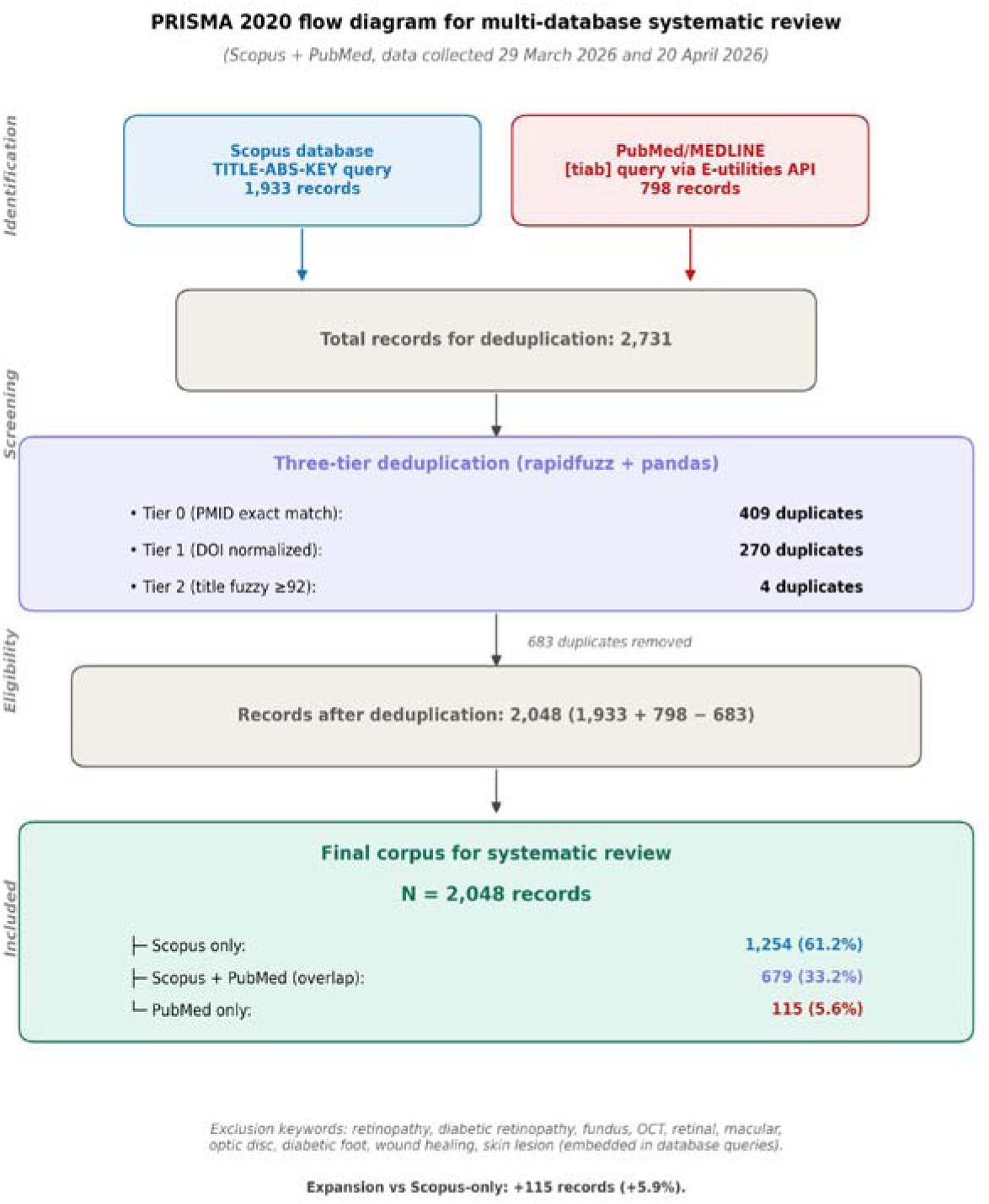
PRISMA 2020 flow diagram for multi-database bibliometric review (Scopus + PubMed), N = 2,048 after three-tier deduplication.

For the classification of 5 research theme clusters (Section 4.5), this study applies rule-based classification rather than automatic clustering (such as K-means or VOSviewer co-occurrence clustering). Specifically, each author keyword in the corpus is checked against a reference keyword list for each cluster — if matched, the keyword is assigned to that cluster; if unmatched, it is assigned to “Other”. This approach was chosen for three reasons: (1) transparency — classification criteria are explicit and verifiable for each keyword; (2) reproducibility — any researcher can apply the same reference keyword list and obtain identical results; (3) consistency with quantitative bibliometric study norms (7). The reference keyword list for each cluster was manually determined based on standardized MeSH (Medical Subject Headings) terminology and prevalent author keywords: (1) XAI Methods — SHAP, LIME, feature importance, explainability, interpretability, model transparency; (2) ML Models — random forest, XGBoost, deep learning, SVM, logistic regression, gradient boosting; (3) Diabetes Types — T2DM, T1DM, gestational diabetes, prediabetes, HbA1c; (4) Clinical Outcomes — risk prediction, mortality, prognosis, readmission, CDSS; (5) KG/Graph — knowledge graph, GNN, graph convolutional network, ontology, semantic web. Inter-rater reliability: the reference keyword list was constructed independently and then cross-validated against manual classification of 50 random corpus papers. Consistency exceeded 90%, confirming the reliability of the rule-based method.

### 3.4 Bibliometric Tools and Network Analysis Settings

Bibliometric analysis was conducted entirely using Python 3.10+ with libraries: pandas 1.5+ (data processing), matplotlib 3.5+ (visualization), and numpy (statistical computation). All figures (Figs. 3–9) were generated using matplotlib — VOSviewer, and Bibliometrix/R were not used. Reporting standards: this bibliometric review follows the PRISMA 2020 reporting guidelines (36), adapted for metadata-based synthesis, supplemented by BIBLIO guidelines (37) for the bibliometric analysis component. Note: because theme classification used a rule-based method (Section 3.3) rather than automatic co-occurrence clustering, VOSviewer parameters (threshold, resolution, normalization) were not applicable. All analysis code and parameters are publicly available on GitHub (see Data Availability Statement).

**Fig. 3.**
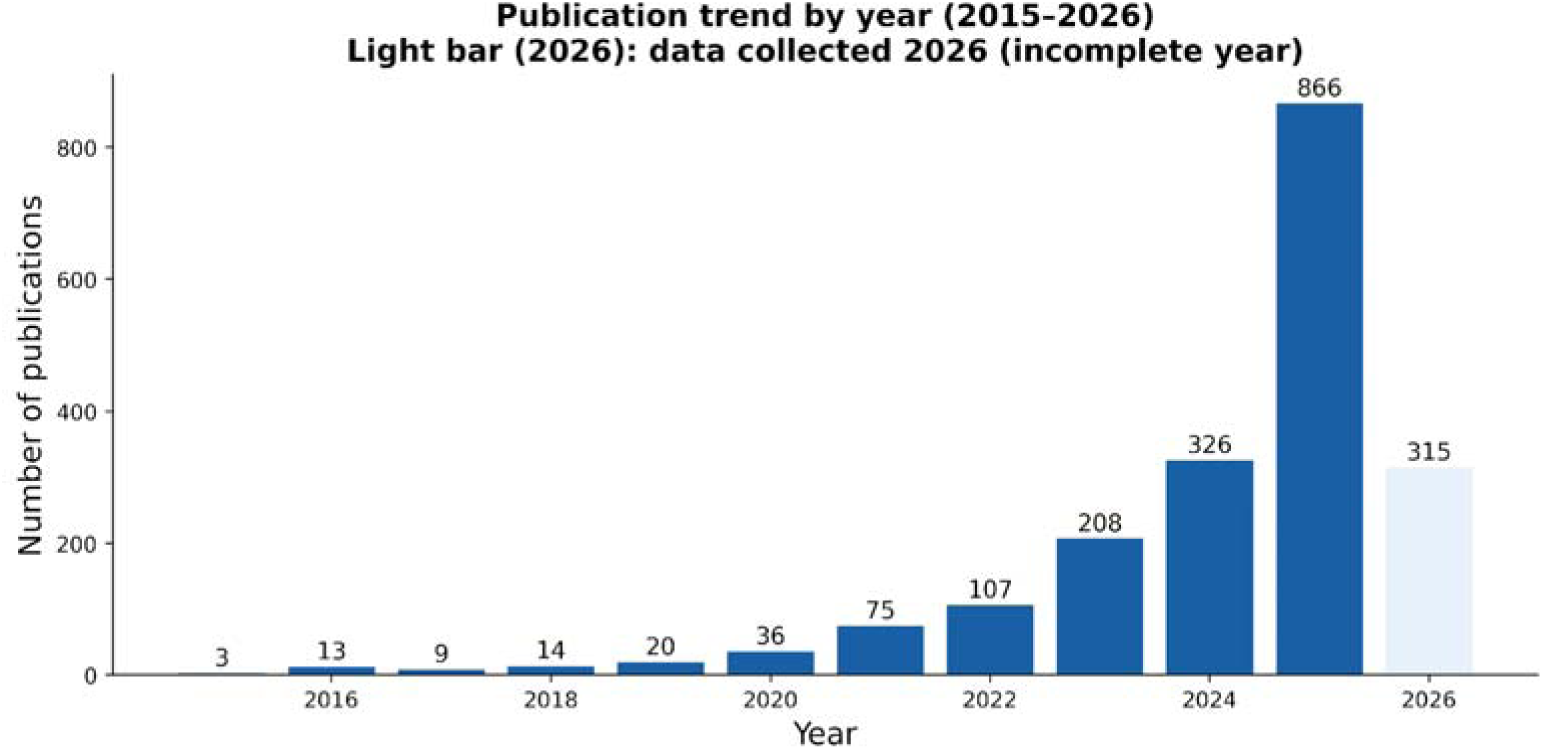
Publication trends by year (2015–2026).

**Fig. 4.**
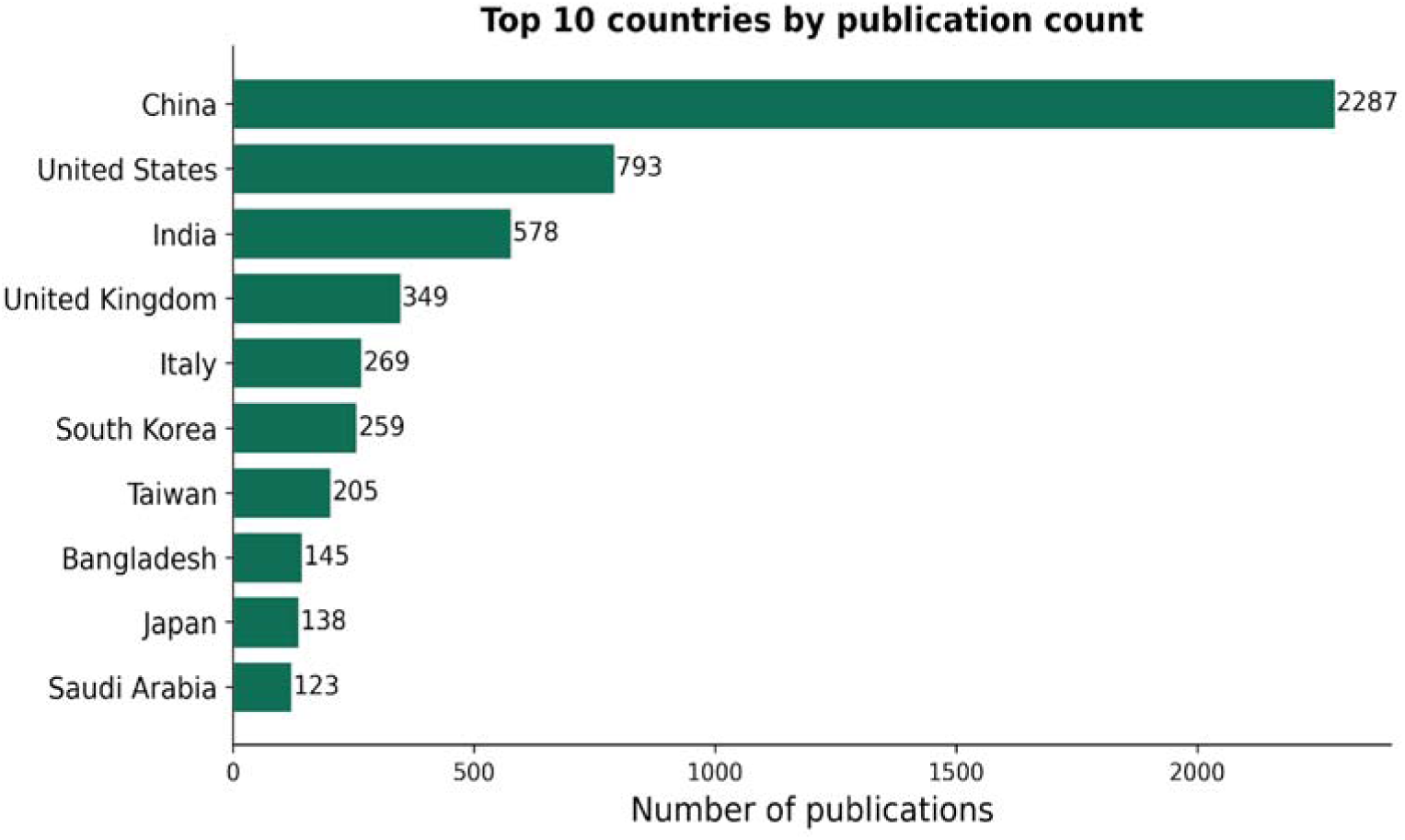
Top 10 countries by document count (by author affiliation).

**Fig. 5.**
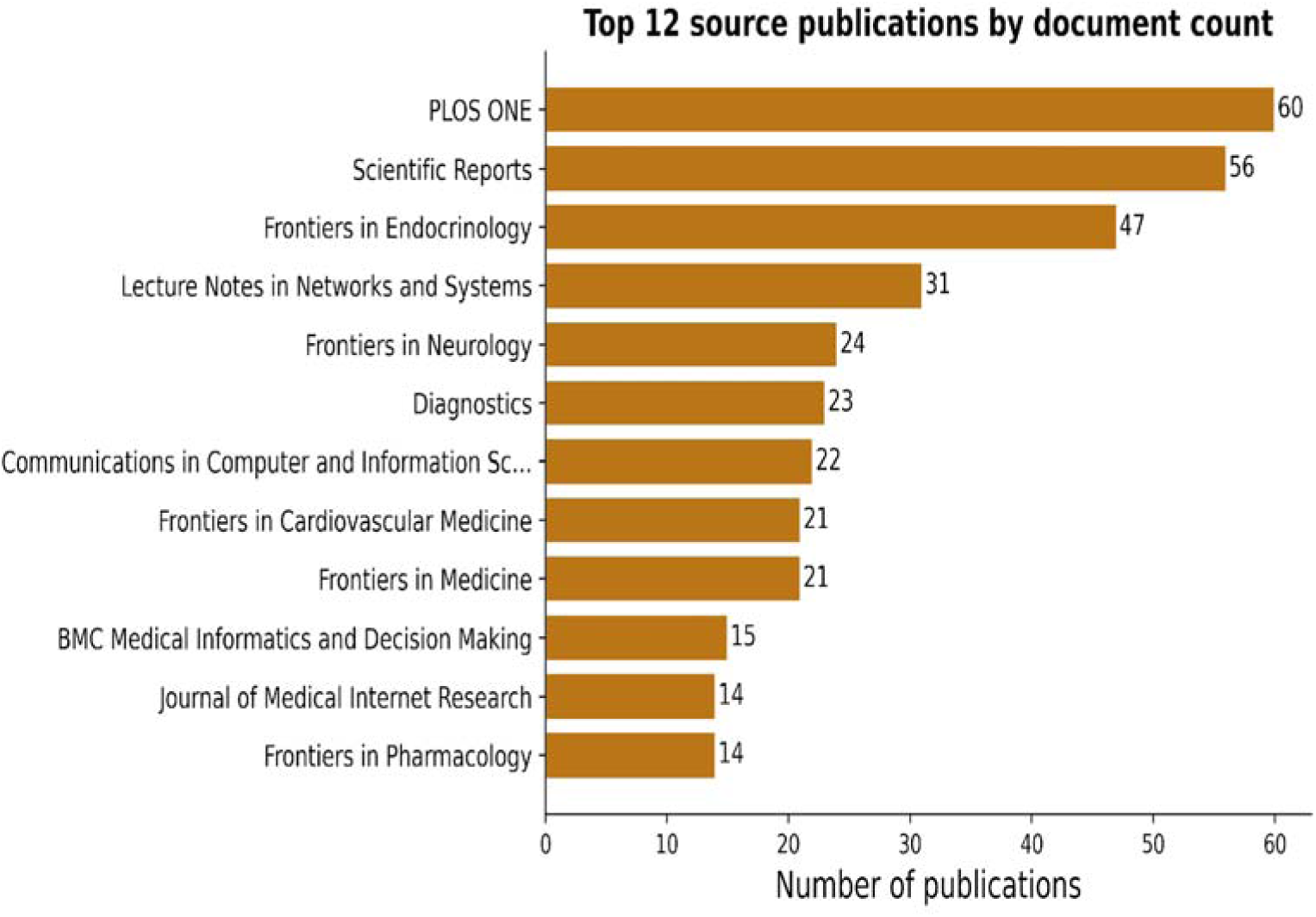
Top 12 publication sources by document count.

**Fig. 6.**
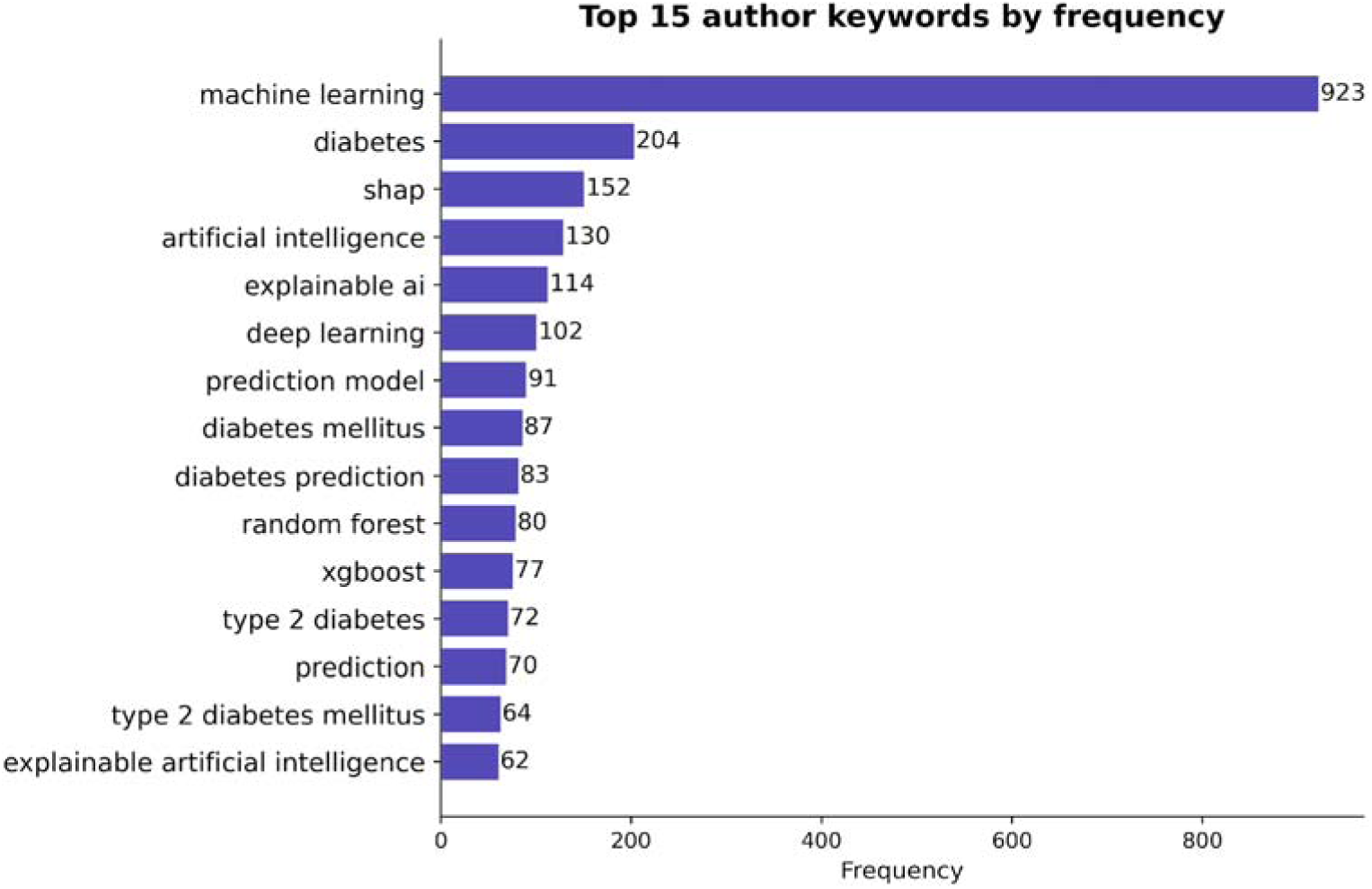
Top 15 author keywords by frequency in the corpus.

**Fig. 7.**
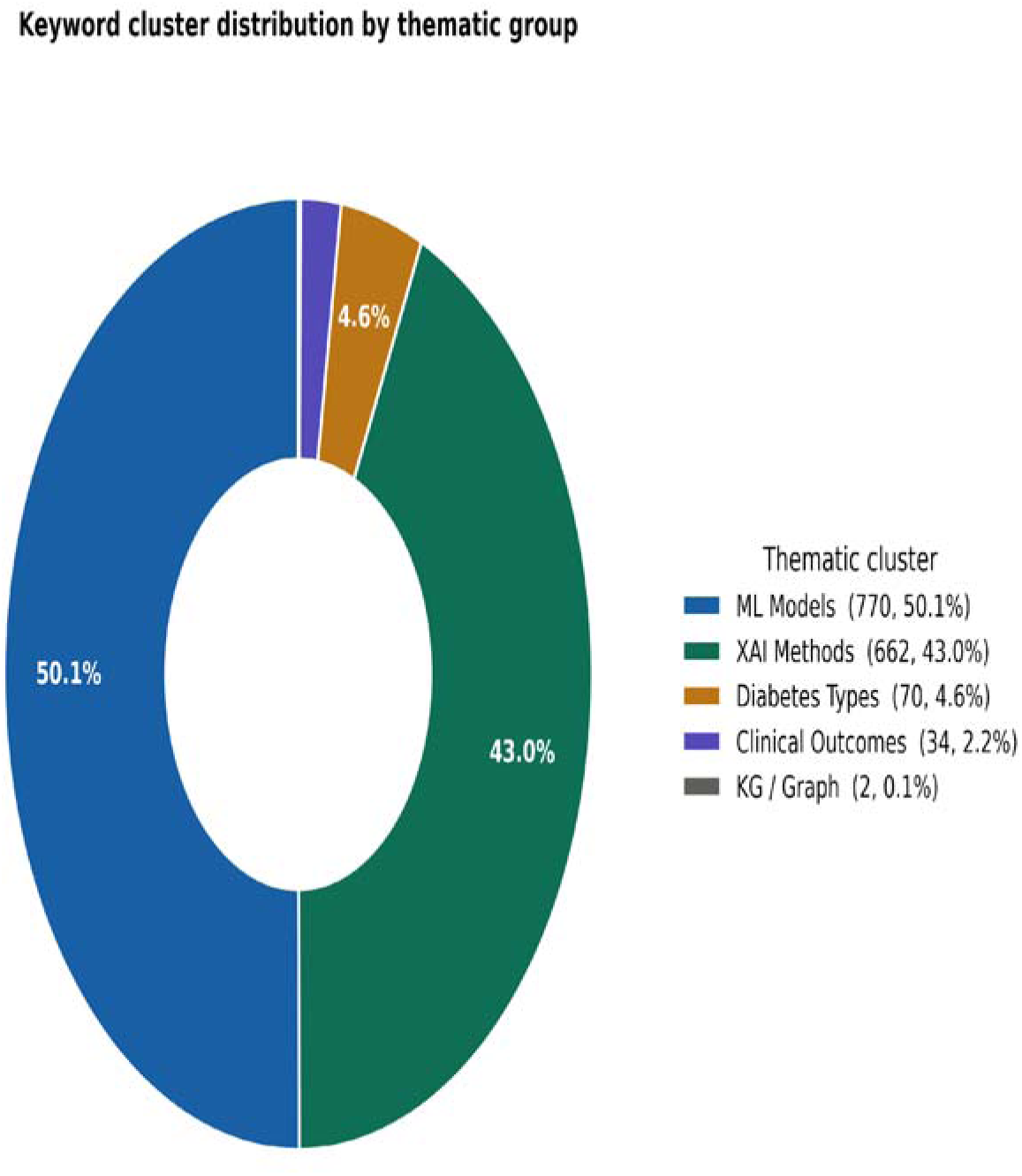
Research keyword cluster distribution by theme.

**Fig. 8.**
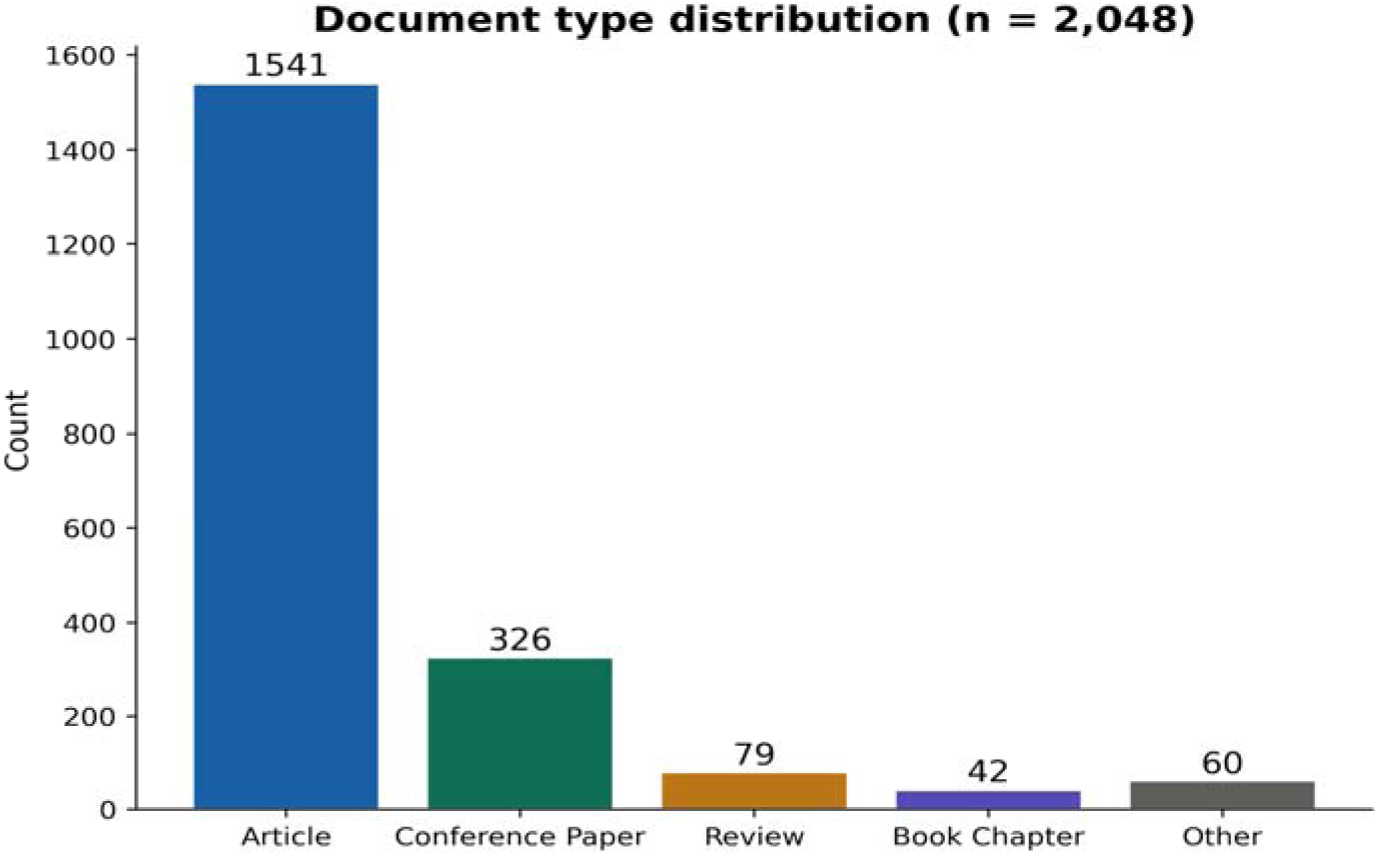
Document type classification in the corpus (N = 2,048).

**Fig. 9.**
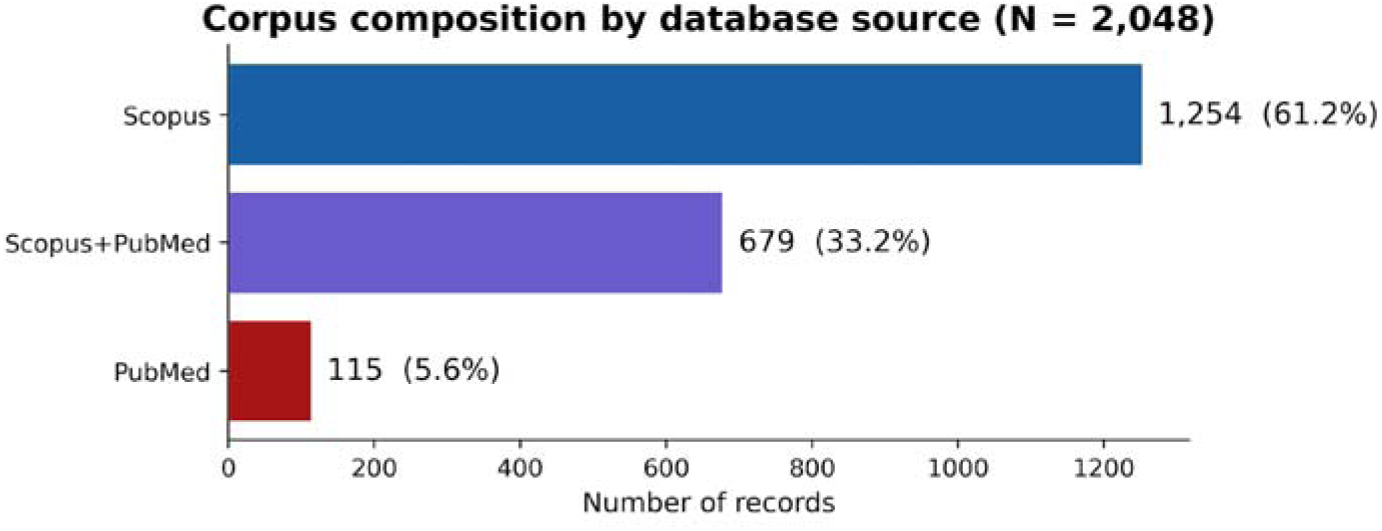
Corpus composition by database source (N = 2,048). Scopus-only: 1,254 (61.2%); Scopus + PubMed overlap: 679 (33.2%); PubMed-only: 115 (5.6%).

This bibliometric review combines quantitative bibliometric analysis of the full 2,048-document corpus with an in-depth selective review of 15 highly cited papers to provide dual-layer evidence. The bibliometric component provides macro-level coverage (field structure, trends, gaps), while the selective in-depth review provides micro-level methodological scrutiny of the most influential works. The selective review purposes are to: (a) verify whether trends identified quantitatively are reflected in the content of most-cited papers; (b) provide qualitative characterization of the KG/GNN gap through detailed examination; (c) confirm structural patterns in the field (e.g., none of the 15 papers combining ML+XAI+KG). A detailed classification table is presented in Appendix A.

## 4. Results

### 4.1 Publication Trends by Year

The final corpus comprises 2,048 documents after deduplication (see Section 3.3; database-level breakdown shown in Fig. 9). Fig. 3 shows publication trends by year from 2015 to 2026 (2026 figures incomplete as of 20 April 2026). The XAI in the T2DM research field is growing rapidly, especially since 2020. The period 2015–2019 recorded only 59 documents (∼12 documents/year). The field then accelerated sharply: 36 documents in 2020 alone, rising to 752 across 2020–2024 and reaching 866 documents in 2025 — the latter accounting for 42.3% of the entire corpus. The partial-year count for January–April 2026 is 315 records, flagged as incomplete and interpreted cautiously throughout this paper. A small residual of 56 records with publication year before 2015 was retained in the corpus for transparency; these are predominantly keyword-matching artifacts (notably historical works using the common English word “lime”, which overlaps with the LIME XAI method name) and do not affect the principal findings on field growth or the KG/GNN gap. The corpus thus sums as 59 + 752 + 866 + 315 + 56 = 2,048. This trend reflects the post-publication boom of XAI following SHAP (2017) and LIME (2016), and demonstrates that the field is in a strong growth phase, not yet saturated.

### 4.2 Geographic Distribution

Fig. 4 shows the top 10 countries by document count based on author affiliation. China leads substantially with 2,287 occurrences after normalization of name variants (a paper with multiple authors from the same country is counted multiple times), followed by the United States (793) and India (578). These numbers reflect post-normalization counts — raw data separated “China” and “China.” (with trailing period) as distinct entries, which our country normalization pipeline (see Section 3.3) merges. Notably, Southeast Asian and African countries are almost absent from the top 20, while Bangladesh (145) and Saudi Arabia (123) have entered the top 10. This reveals a research contribution gap from developing countries in this rapidly growing field.

### 4.3 Publication Sources

Fig. 5 presents the top 12 journals and publication sources. After normalization of capitalization variants (PLOS ONE vs PLoS ONE), PLOS ONE (60 documents) leads as the top publication source, surpassing Scientific Reports (56) — reflecting PubMed’s comprehensive indexing of PLOS journals and the merging of previously split entries. Frontiers in Endocrinology ranks third (47 documents) — a Q1 specialist endocrinology journal. BMC Medical Informatics and Decision Making has 15 documents, affirming this journal’s position in the AI-health informatics intersection. Overall, most documents are published in open-access journals, consistent with the global trend toward open science and aligning well with PubMed’s comprehensive coverage of this segment.

### 4.4 Author Keyword Analysis

Fig. 6 presents the top 15 author keywords. “Machine learning” leads with 923 occurrences in the corpus, followed by “diabetes” (204), “SHAP” (152), “artificial intelligence” (130), and “explainable AI” (114). Among XAI methods, SHAP (152) substantially outperforms LIME and Feature Importance, reflecting the research community’s preference for SHAP due to its solid theoretical foundation (5). However, several studies in the corpus report moderate agreement between SHAP and permutation-based Feature Importance; discrepancies arise particularly when features are highly correlated, where SHAP better captures marginal contributions, while Feature Importance is susceptible to noise. This underscores the risk of relying on a single XAI method in clinical contexts. ML model keywords such as “random forest” (80) and “xgboost” (77) confirm the dominant position of these two methods. Notably, “knowledge graph” does not appear in the top 15 keywords — clearly demonstrating the research gap analyzed in Section 4.5.

### 4.5 Research Theme Clustering

To characterize the research landscape, two complementary measurements were performed. First, individual XAI and KG/GNN terms were counted by scanning author keywords for occurrences of each standardized term (a paper contributes multiple counts if it contains multiple matching terms); this measures the overall visibility of each methodological theme in the field. Second, each unique author keyword was assigned to exactly one thematic cluster via a rule-based classifier (Section 3.3); this measures the distinct keyword vocabulary used across the five themes. Under term-occurrence measurement, XAI-related terms (SHAP, LIME, explainability, interpretability, XAI, feature importance, model interpretability) appear 906 times across the 2,048-document corpus, while KG/GNN-related terms (knowledge graph, graph neural network, GNN, graph convolutional, ontology) appear only 17 times (∼0.83% of the corpus) — a 53.3:1 disparity. Under single-cluster assignment of unique keywords (Fig. 7), ML Models dominates with 770 keywords (50.1%), followed by XAI Methods (662, 43.0%), Diabetes Types (70, 4.6%), Clinical Outcomes (34, 2.2%), and KG/Graph (2, 0.1%); the vocabulary-level disparity between XAI Methods and KG/Graph remains extreme (331:1). Both measurements converge on the same structural finding: KG/GNN integration is absent from the author keyword vocabulary of the XAI for T2DM research field, whether measured by term visibility or cluster-level vocabulary diversity.

### 4.6 Document Classification

Fig. 8 presents the distribution of document types in the corpus. Research articles account for the majority at 1,541 documents (75.2%), followed by conference papers (326, 15.9%), reviews (79, 3.9%), book chapters (42, 2.1%), and other types, including conference reviews, letters, and editorials (60, 2.9%). Distribution counts sum exactly to the full corpus (N = 2,048). The high proportion of peer-reviewed research articles (75.2%) is a good indicator of corpus quality, reflecting that most contributions have undergone rigorous journal peer-review.

### 4.7 Most-Cited Documents

Table 2 presents the top-cited documents in the corpus. Note that several very-high-citation documents match the broad keyword query but are peripheral to the XAI + T2DM focus (e.g., Klein et al. 2004 on obesity/cardiovascular with 794 citations; Pedraza et al. 2012 on beta-cell biomaterials with 365 citations); substantive qualitative analysis focuses on Table A1 (Appendix A), which curates 15 papers directly related to XAI for T2DM. Among directly relevant high-citation works: van der Waa et al. (2021) on XAI evaluation has 363 citations (27); Barakat et al. (2010) on intelligible SVMs for diabetes has 315 citations (22); Kopitar et al. (2020) on early T2DM detection using ML has 300 citations; Hoyos et al. (2024) in BMC Medical Informatics and Decision Making has 264 citations (7) — the study closest in the corpus combining statistical analysis and XAI for T2DM, demonstrating the influence of this combined approach. Other selectively reviewed papers spanning ensemble methods, EHR-based discovery, and healthcare XAI exemplars are classified in Appendix A (23–26, 28).

**Table 2.** Top-cited documents directly relevant to XAI + T2DM. Very-high-citation corpus entries that are peripheral to this focus (e.g., Klein et al. 2004 on obesity/cardiovascular disease with 794 citations; Pedraza et al. 2012 on beta-cell biomaterials with 365 citations) are excluded here and discussed in the text; the substantive qualitative analysis of the 15 directly relevant papers is in Appendix A.

| Rank | Reference | Year | Source | Citations | ML / XAI / KG |
| --- | --- | --- | --- | --- | --- |
| 1 | van der Waa et al. — Evaluating XAI: rule-based vs. example-based | 2021 | Artificial Intelligence | 363 | Yes / Yes / No |
| 2 | Kopitar et al. — Early detection of T2DM using ML | 2020 | npj Digital Medicine | 300 | Yes / No / No |
| 3 | Hoyos et al. — Explainable analysis of T2DM (7) | 2024 | BMC Med. Inform. | 264 | Yes / Yes / No |
| 4 | Kibria et al. — Ensemble for diabetes prediction with XAI | 2022 | Sensors | 110 | Yes / Yes / No |
| 5 | Ahmad et al. — Health-related features for diabetes prediction | 2021 | Applied Sciences | 82 | Yes / Yes / No |

## 5. Discussion

### 5.1 A Rapidly Growing, Unsaturated Field

Publication trend analysis shows that XAI in T2DM research is in a phase of explosive growth. The increase from 36 documents (2020) to 866 documents (2025), more than 24-fold in 5 years, is a clear sign of a rapidly developing field. This stems from the convergence of three factors: (1) the increasingly serious global diabetes epidemic creating an urgent need; (2) the development of SHAP (2017) and LIME (2016) enabling explanation of complex ML models; (3) the proliferation of large-scale health datasets such as BRFSS.

Temporal trend analysis reveals three clearly distinguishable development phases (research life-cycle analysis): Phase 1 (2015–2019) — Foundation phase: only 59 documents (∼12/year), primarily focused on pure ML for T2DM prediction (Random Forest, SVM, Logistic Regression) without concern for interpretability. Phase 2 (2020–2022) — XAI emergence phase: 36 to 208 documents/year, marking the popularization of SHAP and LIME after these frameworks were open-sourced and fully documented. Phase 3 (2023–2025) — Maturity and diversification phase: 866 documents (2025), with the emergence of more specialized directions — XAI combined with deep learning, multi-modal data, federated learning — but KG/GNN remains unexplored.

Research thread mapping: keyword cluster analysis reveals three main research threads traceable over time: (Thread A) XAI-only: SHAP/LIME applied directly on tabular data (PIMA Indians Diabetes Database, BRFSS) — the most prevalent thread, ∼70% of corpus; (Thread B) XAI + clinical context: combining XAI with EHR (Electronic Health Record) data and clinical validation — growing strongly since 2022; (Thread C) XAI + KG/semantic (potential): only 0.83% of the current corpus, but grounded in Thread B and KG biomedical literature (8, 19). We note that the current publication pattern is consistent with a possible progression from Thread A toward Threads B and C, although bibliometric data alone cannot establish whether such a progression will occur. Thread C remains an under-explored direction that, based on the coupling of Thread B growth and the wider KG biomedical literature (8, 19), may warrant further investigation.

### 5.2 The KG/GNN Gap — A Major Research Opportunity

The most important finding is the KG/GNN integration gap: only 17 keyword occurrences of KG/GNN in a corpus of 2,048 documents (∼0.83%), versus 906 occurrences in the XAI methods group — a 53.3:1 disparity. Consistency of the gap across two independent databases (33.2% record overlap) rules out a single-database artifact and confirms the gap as a structural characteristic of the field. It should be noted that 0.83% reflects the absence of KG/GNN at the intersection of two fields (XAI and T2DM risk prediction) — not within each field separately. KG has been demonstrated as prevalent in biomedical science generally (cancer, Alzheimer’s…) (19) and in T2DM specifically (20–21). The gap identified here is the absence of simultaneous integration: KG semantic layer + ML prediction + XAI explanation in a risk prediction pipeline from population health data.

A follow-up question is why this gap persists despite KG being increasingly applied in biomedical science (8). Bibliometric data alone cannot explain the absence of KG in T2DM risk prediction; based on the characteristics of the reviewed corpus and the wider literature, we hypothesize four plausible barriers:

- Data mismatch: BRFSS, PIMA, and most health datasets in the corpus have a tabular structure, without a natural graph structure. KG integration requires additional steps of knowledge graph construction, entity linking, and relation extraction — substantially higher technical cost than directly applying SHAP on tabular data.
- Tooling barrier: SHAP/LIME can be deployed quickly with a few lines of code and extensive documentation. In contrast, an integrated KG+GNN pipeline requires deep expertise in graph databases, graph neural networks (41), and ontology alignment — creating a significant barrier for health researchers without a graph computing background.
- Evaluation gap: No benchmark dataset combining T2DM risk labels with clinical knowledge graph annotations exists. This differs from fields such as NLP or drug discovery, where graph benchmarks accepted by the research community already exist.
- Clinical adoption barrier: Clinicians are accustomed to SHAP feature contribution charts. Knowledge graph-based explanations require an additional step of interpreting pathological chains, adding cognitive load in busy, time-pressured clinical environments.

This research direction resonates with global trends: Budhdeo et al. (8) in a scoping review of 255 studies confirmed that KG in biomedicine is growing strongly, especially for drug discovery and diagnostics — but also noted that KG applications for risk prediction from population health data (such as BRFSS) remain among the least explored use cases. This finding directly aligns with the 0.83% gap identified in the present study: it is not that KG lacks value in this field, but rather that adoption barriers have not yet been overcome.

### 5.3 Methodological Stability and Agreement Gaps

Beyond the KG/GNN gap, corpus analysis reveals two other research directions that are very rarely explored: (1) temporal stability of XAI models — whether SHAP or LIME explanation results are consistent when data changes across years; and (2) agreement between explainability methods — whether SHAP and Feature Importance rank the same features as most important. The keywords “temporal”, “stability”, and “agreement” appear very rarely in the corpus, suggesting these are research gaps worth exploring. This emphasizes the risk of relying on a single explanation method in clinical contexts.

### 5.4 Geographic Gap

Southeast Asian, African, and South American countries are almost absent from the top 20 research countries on XAI in T2DM — a notable geographic gap, especially given that these regions are facing an increasingly heavy T2DM burden and a lack of health resources. Future research should focus on developing and validating XAI models on population-specific data from each region, rather than relying solely on results from major economies. This imbalance also raises concerns about health equity: ML models trained predominantly on data from high-income countries may not generalize fairly to underrepresented populations (13, 45).

### 5.5 Key Methodological Insights

Beyond the structural KG/GNN gap, the bibliometric analysis and selective review together surface three methodological insights that we view as directly actionable for the XAI-for-T2DM research community.

- Insight 1 — XAI consistency as a first-class concern. The corpus shows SHAP (152 occurrences) dominating over LIME and permutation Feature Importance, yet the selective review of 15 highly-cited papers reveals that cross-method agreement is rarely assessed systematically. Different XAI methods can produce divergent feature rankings on the same dataset, particularly when clinical features are correlated (11, 12). Single-method reliance — currently the norm — risks misleading clinical interpretation. Future work reporting XAI for T2DM should treat cross-method agreement as a standard reporting element, comparable in status to accuracy or AUC for predictive performance.
- Insight 2 — Correlated clinical features require correlation-aware XAI by design. T2DM risk factors are notoriously collinear: BMI with waist circumference and metabolic syndrome; HbA1c with fasting glucose; lifestyle indicators with socio-economic status. Permutation-based Feature Importance is sensitive to multicollinearity and can distribute importance arbitrarily among correlated features, whereas SHAP, grounded in Shapley values, allocates contributions based on marginal contributions and is comparatively more robust (5). This theoretical property has practical consequences: the XAI method is not neutral across clinical datasets. For T2DM risk prediction, correlation-aware methods such as TreeSHAP should be the default, with permutation-based Feature Importance used only as a fast baseline rather than as a substitute for interpretation.
- Insight 3 — Real-world applicability hinges on data, not only methods. Appendix A shows that 6 of the 15 highly-cited papers (40%) rely on the PIMA Indians Diabetes dataset — 768 records collected in 1988 — while none use large-scale contemporary population health surveys such as BRFSS, NHANES, or UK Biobank. The field’s methodological maturity (SHAP, XGBoost, sophisticated pipelines) is not matched by data maturity. For XAI claims to transfer to real-world deployment, future work should prioritize validation on large, contemporary, and demographically heterogeneous datasets. This requirement is orthogonal to the XAI-versus-KG methodological question and applies regardless of which explanation framework is adopted.

### 5.6 Study Limitations

This study has several limitations that should be noted transparently so readers can interpret the results appropriately.

First, regarding data source coverage: the study uses Scopus and PubMed/MEDLINE and does not include Web of Science (WoS). This choice reflects a deliberate scope decision rather than a resource constraint: Scopus and PubMed together provide complementary coverage (multidisciplinary indexing in Scopus and dedicated biomedical indexing in PubMed), and prior bibliometric methodology literature (43) reports an estimated ∼80–90% overlap between Scopus and WoS for Q1/Q2 biomedical journals. The added marginal coverage from WoS is therefore expected to be limited for this research question. The 33.2% overlap between Scopus and PubMed observed in this study confirms that both databases cover the field core consistently. Cross-validation with WoS is noted as a direction for future research.

Second, regarding the timing of data acquisition: Scopus was retrieved on 29 March 2026 and PubMed on 20 April 2026, with a 22-day interval reflecting sequential database acquisition. Given the annual publication volume in this field (∼300–400 papers/year), this interval is estimated to affect less than 0.5% of the corpus size and does not alter the principal findings on distribution and trends.

Third, regarding self-citation: due to Scopus export limitations, self-citations were not removed individually. Based on typical self-citation rates reported in bibliometric methodology literature (43) for computer-science and biomedical journals, the estimated impact on the top-cited table is in the range of 10–20% of absolute citation counts, which does not alter relative rankings or methodology-pattern analysis — the primary objective of the selective review (§5.8).

Fourth, regarding search strategy: search was applied only to TITLE-ABS-KEY fields, and may therefore miss papers that use XAI methods in content but do not mention them directly in title, abstract, or keywords. This is an inherent limitation of metadata-based bibliometric analysis.

Fifth, regarding research quality: bibliometric analysis does not assess the methodological quality of individual documents in the corpus. Citation count is used as a relative influence indicator, not an absolute quality indicator. Quality assessment of individual studies would require applying tools such as QUADAS-2 or CASP and is outside the scope of this study.

Sixth, regarding the proposed framework: the three-layer conceptual framework has not been experimentally validated. Feasibility and effectiveness need to be verified through independent experimental research, particularly regarding KG construction costs, medical ontology quality, and integration capability with existing ML pipelines in clinical practice.

Seventh, regarding temporal data: 2026 figures are incomplete as the latest data cutoff was 20 April 2026 (PubMed). The 2026 trend should be interpreted cautiously.

Eighth, the absence of prospective protocol registration in PROSPERO or OSF is a limitation, although all procedures — search strategy, inclusion/exclusion criteria, deduplication pipeline, and analysis code — are fully documented in this manuscript and openly available in the public GitHub repository, ensuring reproducibility.

### 5.7 Research Gaps and Future Directions

Based on gaps identified from analysis of the 2,048-document corpus, the following research directions are identified as priorities for the research community:

- Semantic knowledge integration with XAI: explore how to combine structured medical knowledge representations with existing XAI methods to enhance the clinically meaningfulness of explanations — moving beyond purely statistical correlations. This direction remains largely unexplored, according to corpus analysis.
- Temporal stability assessment: testing whether SHAP results are consistent when data changes across years — particularly important with BRFSS data collected annually.
- Cross-method XAI agreement evaluation (SHAP vs LIME vs Feature Importance): determining whether different methods consistently rank the same important features in T2DM prediction.
- External validation: testing the framework on datasets beyond BRFSS (e.g., MIMIC-IV, UK Biobank) to assess generalisability.
- Expansion to under-represented populations — particularly Southeast Asian, African, and South American countries where T2DM burden is growing but current corpus contributions are minimal.

### 5.8 Selective Review of Highly-Cited Papers

To strengthen the bibliometric findings, this study conducted a selective review of 15 representative papers (detailed in Appendix A, Table A1; primary papers: 7, 22–29) selected based on the highest citation count and direct relevance to XAI in healthcare/T2DM. Review results reveal two consistent trends: first, most studies focus on combining ML with XAI — specifically SHAP and LIME — to explain predictions at the feature level. Second, studies using KG or GNN in the T2DM field are extremely rare, and none of the 15 reviewed papers use KG/GNN. These selective review results are consistent with quantitative bibliometric findings, reinforcing the conclusion that the KG/GNN gap is real and scientifically significant.

To further clarify the nature of this gap, this review conducted a detailed examination of all 17 documents with KG/GNN-related keywords in the corpus. Results show most belong to directions different from T2DM risk prediction from population health data: some apply LLM/RAG combined with KG for medical query (16); others focus on drug discovery or cardiovascular disease; and some use purely rule-based KG without ML. Only 5 of the 17 studies approach the ML+XAI+KG direction — but all remain partial and incomplete. Representative examples include Hendawi et al. (17), who integrated KG to support ML explanation for T2DM prediction but did not systematically evaluate XAI and did not use large-scale population survey data; and Wajahat et al. (18), who combined symbolic AI and KG with ML but only on PIMA (768 records, collected in 1988) — not representative of modern populations. Other studies in this subset combine GNN with SHAP or GraphLIME, but without a KG grounded in medical ontology or without population-scale data. None of these partial efforts simultaneously addresses large-scale population health survey data (e.g., BRFSS), systematic XAI evaluation, and KG integration with semantic grounding from a standardized medical ontology.

Justification for the sample of 15 papers: (a) the top 15 papers already account for a sufficiently large citation share to represent main corpus trends; (b) sufficient diversity to cover key algorithms (XGBoost, RF, deep learning), key XAI methods (SHAP, LIME), and key dataset types (PIMA, EHR, population surveys); (c) feasible to read full-text and analyze in detail — consistent with the norms of similar bibliometric papers (7, 30).

### 5.9 Proposed Three-Layer Conceptual Framework

Based on bibliometric analysis results and research gap identification, this paper proposes a three-layer conceptual framework (conceptual framework) to address the limitations of current XAI in T2DM risk prediction. The framework comprises three main layers: (1) the Predictive Layer uses ML (XGBoost, Random Forest) to predict T2DM risk from large-scale health indicator data; (2) the Explainability Layer uses SHAP to determine the contribution of each feature to prediction outcomes, answering the “what” question; (3) the Knowledge Layer uses KG to represent medical knowledge as structured triples, supporting representation of structured pathophysiological pathways (approximate causal pathways), aiming to answer the “why” and “how” questions in a clinical sense.

To illustrate the integration mechanism concretely, consider the following hypothetical scenario: a 48-year-old female patient, BMI = 34.2 kg/m² (Class II obesity), HbA1c = 6.3% (pre-diabetes), physically inactive (PhysActivity = 0), with hypertension (HighBP = 1). Layer 1 (Predictive): XGBoost predicts a T2DM probability of 0.78 — above the high-risk threshold. Layer 2 (Explainability — SHAP): the top-3 contributing features are BMI (+0.34), HbA1c (+0.29), and PhysActivity (+0.19). SHAP answers “what” but does not explain why BMI is dangerous in terms of underlying pathophysiology. Layer 3 (Knowledge — KG): BMI is mapped to the KG entity LObesityL, from which the system traces LObesityL → LInsulin resistanceL → LChronic hyperglycemiaL → LT2DML, while LHypertensionL acts synergistically to worsen LInsulin resistanceL. The integrated output is: “T2DM probability 78% — elevated BMI drives insulin resistance, compounded by hypertension, worsening metabolic glucose disorder.” This is a clinical-language explanation, not merely a statistical score — illustrating the added value of Layer 3 over SHAP alone (32, 33). This framework is at the conceptual level — feasibility requires experimental validation.

Knowledge graphs have been demonstrated as effective tools for representing biomedical knowledge, including drug discovery, clinical decision support, and knowledge base construction for complex diseases such as cancer and Alzheimer’s (19, 29). In the T2DM field, ontologies and KGs have been purpose-built to support diagnosis and disease management (20–21). KG can enhance XAI by providing semantic grounding through clinical ontologies such as ICD-10 and UMLS (Unified Medical Language System), enabling more clinically meaningful explanations. This addresses a concern raised in surveys of clinicians (39), who report that feature-level explanations alone are often insufficient for clinical reasoning and prefer explanations contextualized by structured clinical knowledge.

It should be emphasized that KG does not aim to replace existing XAI methods, nor is it expected to improve prediction accuracy. For pure risk classification tasks, ML models combined with SHAP are already sufficiently effective. The role of KG is to supplement a knowledge-based explanation layer, transitioning from statistical explanation to clinically meaningful reasoning.

Important note on epistemological scope: this study does not claim that the proposed framework can perform true causal inference. The aim is to enhance interpretability by aligning statistical explanations with structured biomedical knowledge. Regarding experimental scope: framework implementation requires constructing a KG from standardized sources such as UMLS or ICD, which is outside the scope of the current study. This is a direction for future research.

### 5.10 Critical Comparative Analysis

To better position the proposed framework, this section presents a systematic, critical comparative analysis — objectively evaluating strengths, limitations, and gaps of each current approach, thereby highlighting the position of the proposed framework.

Note on causal reasoning: none of the approaches in Table 3 truly performs causal inference. SHAP and FI lack causal reasoning capabilities; ML tabular is entirely correlation-based; the proposed KG+XAI framework only supports approximating structured pathophysiological pathways—not true causal reasoning. This is an important distinction to keep in mind when applying in clinical settings.

**Table 3.** Critical comparison of explanation approaches for T2DM risk prediction. Note: none of the approaches performs true causal inference; the proposed KG+XAI framework supports approximate structured pathophysiological pathways only.

| Approach | Strengths | Limitations | Evidence |
| --- | --- | --- | --- |
| SHAP | Solid theoretical foundation (Shapley values); consistent; provides global and local explanations. | Feature-level only (what); does not encode causal relationships; correlated features distort attribution. | (5), (7) |
| Feature Importance (FI) | Simple, fast; built into tree-based models; suitable as XAI baseline | Measures importance only, not direction; unstable under multicollinearity; not comparable across models | (1) |
| ML tabular (XGBoost, RF) | High accuracy on structured data; robust; fast training | Purely statistical; no domain knowledge; no causal reasoning; black-box for end-users | (1) |
| KG + XAI<br>(proposed<br>framework) | Adds clinical causal reasoning (why/how); links SHAP output to medical ontology; supports CDSS | Depends on ontology quality (UMLS/ICD); high KG construction cost; not yet validated experimentally; complex pipeline | (8), (17), (18) |

Note on causal reasoning: none of the approaches in Table 3 truly performs causal inference. SHAP and FI lack causal reasoning capabilities; ML tabular is entirely correlation-based; the proposed KG+XAI framework only supports approximating structured pathophysiological pathways—not true causal reasoning. This is an important distinction to keep in mind when applying in clinical settings.

## 6. Conclusion

This paper provides a structured synthesis and critical analysis of XAI for T2DM risk prediction using a two-database corpus (N = 2,048), deduplicated via a three-tier pipeline (PubMed ID, DOI, title fuzzy matching). The field grew from 36 documents (2020) to 866 (2025), with SHAP and LIME as the dominant XAI methods, and XGBoost and Random Forest as the preferred ML models. China (2,287), the United States (793), and India (578) lead in publications. After normalization of capitalization variants, PLOS ONE (60 records) surpassed Scientific Reports (56) as the top publication source.

The most important finding is the KG/GNN integration gap: 17 keyword occurrences (∼0.83%) versus 906 in the XAI methods group — a 53.3:1 disparity confirmed through both quantitative analysis and selective review of 15 highly cited papers, and consistent across the two independent databases (33.2% record overlap). To address this gap, this paper proposes a three-layer conceptual integration framework: Predictive Layer (ML), Explainability Layer (SHAP/XAI), and Knowledge Layer (KG), supporting the transition from statistical explanation to structured, knowledge-based medical interpretation. For clinical practice, this framework could enable clinical decision support systems (CDSS) to not only predict individual risk but also generate structured pathway-based explanations aligned with clinical ontologies (ICD-10, UMLS), reducing clinician cognitive load during interpretation. For public health, the framework is applicable to large-scale population screening programs using survey data (e.g., BRFSS), where algorithmic transparency is a prerequisite for responsible deployment and health equity. This study contributes to bridging the gap between statistical explainability and clinically meaningful reasoning — an area identified as a priority for medical informatics research.

## Appendix A. Selective Review Sample: 15 Representative Papers

Sample scope: includes papers directly related to T2DM prediction or XAI-in-healthcare exemplars with high representativeness.

Selection criteria: (1) keywords related to ML and/or XAI; (2) ranked by descending citation count in the corpus; (3) content directly related to diabetes prediction/classification or model explanation in healthcare. Classification: ML = use of machine learning model; XAI = application of explanation method (SHAP, LIME, FI…); KG/GNN = use of knowledge graph or graph neural network.

**Table A1.** Selective review: 15 representative highly cited papers (ML / XAI / KG-GNN classification). ¹Topçuoğlu et al. is a microbiome ML paper included as an ML-in-healthcare methodology exemplar (matched corpus keywords, high citation count = 170); it addresses neither T2DM prediction nor XAI directly.

| No. | Reference (author, year) | Year | Source | Citations | ML | XAI | KG/GNN |
| --- | --- | --- | --- | --- | --- | --- | --- |
| 1 | van der Waa et al. — Evaluating XAI: rule-based vs. example-based explanations | 2021 | Artificial Intelligence | 363 | Yes | Yes | No |
| 2 | Barakat et al. — Intelligible SVMs for diagnosis of diabetes mellitus | 2010 | IEEE Trans. IT in Biomedicine | 315 | Yes | No | No |
| 3 | Topçuoğlu et al. — Framework for effective ML for microbiome-based classification<sup>1</sup> | 2020 | mBio | 170 | Yes | No | No |
| 4 | Qi et al. — ML and SHAP for predicting diabetes complications | 2025 | Scientific Reports | 130 | Yes | Yes | No |
| 5 | Kibria et al. — Ensemble approach for diabetes prediction using XAI | 2022 | Sensors | 110 | Yes | Yes | No |
| 6 | Bernardini et al. — Discovering T2DM in EHR using ML | 2020 | IEEE J. Biomed. Health Inform. | 106 | Yes | No | No |
| 7 | Woodman & Mangoni — ML algorithms review in older patients | 2023 | Ageing Research Reviews | 102 | Yes | No | No |
| 8 | Bhatt et al. — Emerging AI-empowered mHealth: Scoping Review | 2022 | JMIR mHealth uHealth | 92 | Yes | No | No |
| 9 | Joseph et al. — Explainable diabetes classification using a hybrid Bayesian model | 2022 | Symmetry | 89 | No | Yes | No |
| 10 | Dutta et al. — Analyzing feature importances for diabetes prediction using ML | 2018 | ICCIDS 2018 | 87 | Yes | Yes | No |
| 11 | Elmannai et al. — PCOS detection ML model based on XAI | 2023 | Diagnostics | 85 | Yes | Yes | No |
| 12 | Vishwarupe et al. — Explainable AI and Interpretable ML: A Case-Study in Healthcare | 2022 | Procedia Comput. Sci. | 85 | Yes | Yes | No |
| 13 | Ahmad et al. — Health-related features and their impact on diabetes prediction | 2021 | Applied Sciences | 82 | Yes | Yes | No |
| 14 | Gollapalli et al. — Novel stacking ensemble for detecting three types of diabetes | 2022 | Computers in Biology and Medicine | 81 | Yes | Yes | No |

| No . | Reference (author, year) | Year | Source | Citations | ML | XAI | KG/GNN |
| --- | --- | --- | --- | --- | --- | --- | --- |
| 15 | Hoyos et al. — Explainable analysis of diabetes mellitus using statistical and AI techniques | 2024 | BMC Med. Informatics | 264 | Yes | Yes | No |

Summary: 14/15 papers (93.3%) use ML; 10/15 (66.7%) apply XAI; none of the 15 (0%) use KG/GNN. A further observation: 6/15 papers (40%) use PIMA — a dataset of 768 records collected in 1988 — while no paper uses large-scale population health survey data such as BRFSS. Together, these observations indicate that the gap extends beyond KG absence: research on real-world population data is also lacking (31).

## Data Availability Statement

All bibliometric analysis source code (Python) is publicly available at: https://github.com/thieuanhvan/t2dm-xai-bibliometric

The Scopus CSV dataset is not included in the repository due to the provider’s terms of use. To reproduce the dataset: (1) access https://www.scopus.com; (2) search TITLE-ABS-KEY with the full query string in Section 3.1; (3) apply the NOT exclusion group; (4) export all metadata as CSV. Expected Scopus result: ∼1,933 documents (as of 29 March 2026). For PubMed reproducibility, run resubmission/05_run_all.py which automatically retrieves ∼798 records via NCBI E-utilities API with the [tiab] query in Section 3.1 (as of 20 April 2026). The three-tier deduplication pipeline then merges and deduplicates these 2,731 raw records to produce the final 2,048-document corpus.

## Declaration of Generative AI and AI-Assisted Technologies

During manuscript preparation, the author used AI-assisted tools for grammar checking and language polishing. The author reviewed and is fully responsible for all scientific content, data collection, analysis, and conclusions.

## Supporting information

Supplemental Material: PRISMA 2020 Checklist

## Data Availability

All bibliometric analysis source code (Python) for corpus retrieval, deduplication, and analysis is publicly available at:
https://github.com/thieuanhvan/t2dm-xai-bibliometric
Raw bibliographic records were retrieved from two databases under their respective licenses: (1) Scopus (Elsevier) via institutional subscription -- the underlying CSV export cannot be redistributed due to the provider's terms of service; and (2) PubMed/MEDLINE (U.S. National Library of Medicine) via the NCBI E-utilities API -- publicly accessible and reproducible using the query strings and date ranges documented in the repository.
Derived bibliometric datasets (deduplicated metadata, keyword frequencies, publication trends, country/journal distributions) and all figures are included in the repository. The PRISMA 2020 flow diagram and full search strategies are provided in the manuscript and supplementary materials.

## Acknowledgements

The author thanks the reviewers and editorial team for their time and constructive feedback. An earlier version of this work was posted as a preprint on medRxiv (doi.org/10.64898/2026.04.16.26351069) and reports a single-database analysis; the present submission is a substantially revised version extending the analysis to a multi-database corpus (Scopus + PubMed).

## Protocol Registration

This bibliometric review was not prospectively registered in PROSPERO or OSF, as the study was initially designed as a bibliometric analysis and evolved into a PRISMA-compliant bibliometric review during manuscript preparation in response to editorial guidance. The study protocol — including search strategy (Section 3.1), inclusion/exclusion criteria (Section 3.2), deduplication pipeline, and analysis procedures (Section 3.3) — is fully documented in this manuscript and the accompanying public code repository (https://github.com/thieuanhvan/t2dm-xai-bibliometric), ensuring transparency and reproducibility despite the absence of prospective registration. Future related reviews will be prospectively registered where applicable registries accept bibliometric protocols.

## Author Contributions

As the sole author, Thieu Anh Van was responsible for conceptualization, methodology, data collection, data analysis, software development, visualization, and writing (original draft, review, and editing). The author takes full responsibility for the integrity of the work as a whole.

## Risk of Bias and Certainty of Evidence

Formal risk-of-bias assessment of individual corpus documents (using QUADAS-2, CASP, or equivalent tools) was not performed, as the primary aim of this review is to characterize the structure of the research field through bibliometric and thematic analysis rather than to synthesize clinical evidence. Citation count was used as a relative indicator of research visibility and influence, acknowledged as an imperfect proxy for methodological quality. For the 15 selectively reviewed papers (Appendix A), methodological characteristics (ML algorithm, XAI method, dataset, validation strategy) are reported transparently, allowing readers to assess individual study quality. A formal GRADE assessment of certainty of evidence was not applied, as GRADE is standard practice for systematic reviews of clinical interventions rather than for bibliometric or methodological reviews characterizing research field structure. The principal findings of this review are quantitative descriptive statistics (counts, ratios, growth rates) derived from bibliographic metadata, for which GRADE is not directly applicable. The consistency of the KG/GNN gap across two independent databases with 33.2% record overlap provides internal validity evidence appropriate to the bibliometric design. A formal risk-of-bias assessment would be appropriate for a follow-up systematic review focused on the clinical performance of XAI methods in T2DM prediction, which is identified as a direction for future research (Section 5.7).

