## Supplemental Material: PRISMA 2020 Checklist for "A Bibliometric Review of Explainable AI in Diabetes Risk Prediction: Trends, Gaps, and Knowledge Graph Opportunities"

Submission to Frontiers in Digital Health — Health Informatics Section — Systematic Review

Author: Thieu Anh Van	|	University of Information Technology, VNU-HCM	|	21 April 2026

This checklist documents adherence to the **PRISMA 2020 Statement** (Page MJ, McKenzie JE, Bossuyt PM, et al. *The PRISMA 2020 statement: an updated guideline for reporting systematic reviews*. BMJ 2021;372:n71. doi:10.1136/bmj.n71) for the submitted manuscript.

All 27 items of the PRISMA 2020 checklist are addressed. The "Location" column references the corresponding section, subsection, figure, or table of the manuscript (version 82, dated 21 April 2026, 2,048-document two-database corpus: Scopus + PubMed/MEDLINE). The manuscript is classified as a **Systematic Bibliometric Review** — a review type accepted under the Systematic Review article category at Frontiers in Digital Health, which explicitly includes mapping reviews, scoping reviews, and related synthesis formats. Because the synthesis operates on bibliographic metadata rather than clinical-outcome data, PRISMA 2020 is applied with its standard adaptations for metadata-based synthesis. Where an item is not directly applicable to the bibliometric design — for example, meta-analytic effect measures, GRADE certainty assessment for clinical outcomes, or individual-study risk-of-bias tools (QUADAS-2, CASP) — the rationale is provided in the "Risk of Bias and Certainty of Evidence" section of the manuscript and restated here for transparency.

Because this review evolved from a bibliometric analysis to a PRISMA-compliant bibliometric review during manuscript preparation, prospective protocol registration (PROSPERO/OSF) was not undertaken; the protocol is fully documented in Sections 3.1–3.4 of the manuscript and in the public GitHub repository https://github.com/thieuanhvan/t2dm-xai-bibliometric. This limitation is acknowledged in item 24a below.

**PRISMA 2020 Checklist — 27 Items**

| **Section and Topic** | **Item #** | **Checklist item** | **Location where item is reported** |
| --- | --- | --- | --- |
| **TITLE** | | | |
| **Title** | **1** | Identify the report as a systematic review. | Title page (line 1); Article-type banner: "Article Type: Systematic Bibliometric Review" |
| **ABSTRACT** | | | |
| **Abstract** | **2** | See the PRISMA 2020 for Abstracts checklist. Use structured format including Background, Methods, Results, and Conclusions. | Abstract (structured: Background, Methods, Results, Conclusions) |
| **INTRODUCTION** | | | |
| **Rationale** | **3** | Describe the rationale for the review in the context of existing knowledge. | §1.1 Research Motivation; §1.2 Problem Statement and Research Gaps |
| **Objectives** | **4** | Provide an explicit statement of the objective(s) or question(s) the review addresses. | §1.3 Research Questions (RQ1–RQ4); §1.4 Principal Contributions |
| **METHODS** | | | |
| **Eligibility criteria** | **5** | Specify the inclusion and exclusion criteria for the review and how studies were grouped for the syntheses. | §3.2 Inclusion and Exclusion Criteria |
| **Information sources** | **6** | Specify all databases, registers, websites, organisations, reference lists, and other sources searched or consulted to identify studies. Specify the date when each source was last searched or consulted. | §3.1 Data Source and Search Strategy (Scopus, 29 March 2026; PubMed/MEDLINE, 20 April 2026) |
| **Search strategy** | **7** | Present the full search strategies for all databases, registers, and websites, including any filters and limits used. | §3.1 (full Scopus TITLE-ABS-KEY query + equivalent PubMed [tiab] query, including NOT exclusion group). Supplementary Material |
| **Selection process** | **8** | Specify the methods used to decide whether a study met the inclusion criteria of the review, including how many reviewers screened each record and each report retrieved, whether they worked independently, and if applicable, details of automation tools used in the process. | §3.1 (automated keyword-based selection via database queries; single-author workflow documented in §"Author Contributions"); §3.3 (data processing pipeline) |
| **Data collection process** | **9** | Specify the methods used to collect data from reports, including how many reviewers collected data from each report, whether they worked independently, any processes for obtaining or confirming data from study investigators, and if applicable, details of automation tools used in the process. | §3.1 (Scopus CSV export; PubMed via NCBI E-utilities API using Biopython); §3.3 (harmonization to common schema; automated extraction) |
| **Data items** | **10a** | List and define all outcomes for which data were sought. Specify whether all results that were compatible with each outcome measure in each study were sought (e.g. for all measures, time points, analyses), and if not, the methods used to decide which results to collect. | §3.3 (bibliographic metadata fields: title, authors, year, source, DOI, PMID, abstract, keywords, affiliations, citation count) |
| **Data items** | **10b** | List and define all other variables for which data were sought (e.g. participant and intervention characteristics, funding sources). Describe any assumptions made about any missing or unclear information. | §3.3 (synonym merging; compound-term normalization; lowercase conversion); §5.6 (limitations regarding self-citations and missing metadata) |
| **Study risk of bias assessment** | **11** | Specify the methods used to assess risk of bias in the included studies, including details of the tool(s) used, how many reviewers assessed each study and whether they worked independently, and if applicable, details of automation tools used in the process. | §"Risk of Bias and Certainty of Evidence" (formal risk-of-bias assessment not performed; rationale: bibliometric characterization rather than clinical-evidence synthesis; methodological characteristics reported transparently for 15 selectively reviewed papers in Appendix A) |
| **Effect measures** | **12** | Specify for each outcome the effect measure(s) (e.g. risk ratio, mean difference) used in the synthesis or presentation of results. | §3.3, §3.4, §4 (descriptive bibliometric measures: keyword frequency counts, publication counts by year/country/source, citation counts, ratio statistics [e.g., 53.3:1 XAI-to-KG disparity], percentage overlap [33.2%]); not applicable for clinical-effect meta-analysis |
| **Synthesis methods** | **13a** | Describe the processes used to decide which studies were eligible for each synthesis (e.g. tabulating the study intervention characteristics and comparing against the planned groups for each synthesis). | §3.3 (rule-based thematic classification into 5 clusters: XAI Methods, ML Models, Diabetes Types, Clinical Outcomes, KG/Graph); §5.8 (selection of 15 highly cited papers for selective review) |
| **Synthesis methods** | **13b** | Describe any methods required to prepare the data for presentation or synthesis, such as handling of missing summary statistics, or data conversions. | §3.3 (three-tier deduplication pipeline: PubMed ID → normalized DOI → title fuzzy matching; country-name normalization; capitalization-variant merging for source titles) |
| **Synthesis methods** | **13c** | Describe any methods used to tabulate or visually display results of individual studies and syntheses. | §3.4 (Python 3.10+ with pandas 1.5+, matplotlib 3.5+, numpy); Figs. 2–9 (PRISMA flow, temporal trends, geographic distribution, sources, keywords, theme clusters, document types, database-source composition); Tables 1–2; Table A1 |
| **Synthesis methods** | **13d** | Describe any methods used to synthesize results and provide a rationale for the choice(s). If meta-analysis was performed, describe the model(s), method(s) to identify the presence and extent of statistical heterogeneity, and software package(s) used. | §3.3 (rule-based synthesis; rationale: transparency, reproducibility, consistency with bibliometric norms; inter-rater reliability >90% via cross-validation of 50 random corpus papers). Meta-analysis not applicable (bibliometric design) |
| **Synthesis methods** | **13e** | Describe any methods used to explore possible causes of heterogeneity among study results (e.g. subgroup analysis, meta-regression). | §4.5 (dual measurement: term-occurrence count vs. single-cluster assignment of unique keywords to explore robustness across clustering granularity); §4.1 (temporal subgroup analysis: three development phases 2015–2019 / 2020–2022 / 2023–2025) |
| **Synthesis methods** | **13f** | Describe any sensitivity analyses conducted to assess robustness of the synthesized results. | §4.5 (sensitivity to measurement method: both term-occurrence [53.3:1] and vocabulary-level [331:1] measurements converge on the same structural finding); §5.2 (cross-database consistency check: 33.2% overlap rules out single-database artifact) |
| **Reporting bias assessment** | **14** | Describe any methods used to assess risk of bias due to missing results in a synthesis (arising from reporting biases). | §5.6 (multi-database design addresses database-coverage bias; TITLE-ABS-KEY-only search limitation acknowledged); §"Risk of Bias and Certainty of Evidence" (33.2% Scopus–PubMed overlap provides internal-validity evidence) |
| **Certainty assessment** | **15** | Describe any methods used to assess certainty (or confidence) in the body of evidence for an outcome. | §"Risk of Bias and Certainty of Evidence" (formal GRADE not applied; rationale: GRADE is standard for clinical-intervention reviews rather than bibliometric/methodological field-structure reviews; cross-database consistency used as internal-validity evidence) |
| **RESULTS** | | | |
| **Study selection** | **16a** | Describe the results of the search and selection process, from the number of records identified in the search to the number of studies included in the review, ideally using a flow diagram. | §3.1, §4 (Fig. 2 — PRISMA 2020 flow diagram; Scopus 1,933 + PubMed 798 = 2,731 raw → 2,048 after three-tier deduplication); §3.3 |
| **Study selection** | **16b** | Cite studies that might appear to meet the inclusion criteria, but which were excluded, and explain why they were excluded. | §4.7 (high-citation but topically peripheral entries excluded from focused Table 2 analysis: Klein et al. 2004 [794 cites, obesity/CVD]; Pedraza et al. 2012 [365 cites, beta-cell biomaterials]); §3.2 (NOT-group exclusions: retinopathy, diabetic foot, wound healing) |
| **Study characteristics** | **17** | Cite each included study and present its characteristics. | §4 (corpus characteristics: N = 2,048; §4.1 year distribution; §4.2 geography; §4.3 sources; §4.4 keywords; §4.5 theme clusters; §4.6 document types; §4.7 top-cited; Table 2); Appendix A (Table A1 — 15 selectively reviewed papers with author, year, source, citations, ML/XAI/KG classification) |
| **Risk of bias in studies** | **18** | Present assessments of risk of bias for each included study. | §"Risk of Bias and Certainty of Evidence" (individual-study risk-of-bias assessment not performed — rationale stated; methodological characteristics of 15 selectively reviewed papers reported in Appendix A for reader assessment) |
| **Results of individual studies** | **19** | For all outcomes, present, for each study: (a) summary statistics for each group (where appropriate) and (b) an effect estimate and its precision (e.g. confidence/credible interval), ideally using structured tables or plots. | Table 2 (top-cited documents with citations and ML/XAI/KG classification); Table A1 (15 selectively reviewed papers with ML/XAI/KG classification). For bibliometric outcome measures (counts/ratios), confidence intervals not applicable to census-level corpus analysis |
| **Results of syntheses** | **20a** | For each synthesis, briefly summarise the characteristics and risk of bias among contributing studies. | §4.1–4.7 (corpus characteristics by synthesis dimension); §5.8 (selective-review methodology and justification for the 15-paper sample) |
| **Results of syntheses** | **20b** | Present results of all statistical syntheses conducted. If meta-analysis was done, present for each the summary estimate and its precision (e.g. confidence/credible interval) and measures of statistical heterogeneity. If comparing groups, describe the direction of the effect. | §4.1 (temporal growth: 36→866 docs/year, 24× in 5 years); §4.2 (top countries: China 2,287, USA 793, India 578); §4.4–4.5 (XAI 906 vs. KG/GNN 17 = 53.3:1); §4.5 (cluster distribution: ML 50.1%, XAI 43.0%, Diabetes 4.6%, Clinical 2.2%, KG 0.1%); §4.6 (document types); Figs. 3–9 |
| **Results of syntheses** | **20c** | Present results of all investigations of possible causes of heterogeneity among study results. | §4.1 (three temporal phases: Foundation 2015–2019; Emergence 2020–2022; Maturity 2023–2025); §5.1 (research-thread mapping: Thread A / B / C); §4.5 (measurement-method comparison: term-occurrence vs. vocabulary-level) |
| **Results of syntheses** | **20d** | Present results of all sensitivity analyses conducted to assess the robustness of the synthesized results. | §4.5 (vocabulary-level 331:1 vs. term-occurrence 53.3:1 — both confirm KG/GNN absence); §5.2 (cross-database consistency: gap confirmed across Scopus and PubMed with 33.2% overlap) |
| **Reporting biases** | **21** | Present assessments of risk of bias due to missing results (arising from reporting biases) for each synthesis assessed. | §5.6 (database-coverage limitations: WoS not included; 22-day acquisition interval between Scopus and PubMed); §5.6 (search-field limitation: TITLE-ABS-KEY/[tiab] only); §5.6 (self-citations not removed individually — estimated 10–20% with negligible impact on trend analysis) |
| **Certainty of evidence** | **22** | Present assessments of certainty (or confidence) in the body of evidence for each outcome assessed. | §"Risk of Bias and Certainty of Evidence" (formal GRADE not applied; internal validity supported by cross-database consistency — 33.2% overlap with convergent KG/GNN-gap finding; dual confirmation via quantitative bibliometrics and selective review of 15 papers) |
| **DISCUSSION** | | | |
| **Discussion** | **23a** | Provide a general interpretation of the results in the context of other evidence. | §5.1 (unsaturated-field interpretation); §5.2 (KG/GNN gap interpreted in context of Budhdeo et al. scoping review); §5.3 (methodological stability and agreement gaps); §5.5 Key Methodological Insights (XAI consistency, correlated features, real-world applicability); §5.9 (three-layer conceptual framework) |
| **Discussion** | **23b** | Discuss any limitations of the evidence included in the review. | §5.6 (six limitation categories: database coverage; acquisition timing; search strategy; research quality not individually assessed; framework not experimentally validated; 2026 data incomplete; absence of prospective protocol registration) |
| **Discussion** | **23c** | Discuss any limitations of the review processes used. | §5.6 (TITLE-ABS-KEY-only search; metadata-based analysis; self-citations not individually removed; rule-based rather than automatic clustering — trade-offs discussed); §"Protocol Registration" (non-prospective registration) |
| **Discussion** | **23d** | Discuss implications of the results for practice, policy, and future research. | §5.2 (four barriers to KG adoption); §5.5 (three methodological insights: XAI consistency, correlated features, real-world applicability); §5.7 Research Gaps and Future Directions (five priority directions: semantic knowledge integration; temporal stability; cross-method XAI agreement; external validation; under-represented populations); §6 Conclusion (CDSS and population-screening implications) |
| **OTHER INFORMATION** | | | |
| **Registration and protocol** | **24a** | Provide registration information for the review, including the register name and registration number, or state that the review was not registered. | §"Protocol Registration" (not prospectively registered in PROSPERO or OSF; rationale provided) |
| **Registration and protocol** | **24b** | Indicate where the review protocol can be accessed, or state that a protocol was not prepared. | §"Protocol Registration" (full protocol — search strategy §3.1, inclusion/exclusion §3.2, deduplication and analysis §3.3 — documented in manuscript and in public GitHub repository) |
| **Registration and protocol** | **24c** | Describe and explain any amendments to information provided at registration or in the protocol. | §"Protocol Registration" (study evolved from bibliometric analysis to systematic-review format during manuscript preparation; Acknowledgements discloses earlier single-database medRxiv preprint [doi.org/10.64898/2026.04.16.26351069] as the prior version) |
| **Support** | **25** | Describe sources of financial or non-financial support for the review, and the role of the funders or sponsors in the review. | §"Funding" (no specific grant from public, commercial, or not-for-profit sectors; no APC received at time of submission) |
| **Competing interests** | **26** | Declare any competing interests of review authors. | §"Declaration of Competing Interests" (author declares no competing financial or non-financial interests) |
| **Availability of data, code and other materials** | **27** | Report which of the following are publicly available and where they can be found: template data collection forms; data extracted from included studies; data used for all analyses; analytical code; any other materials used in the review. | §"Data Availability Statement" (all Python analysis code publicly available at https://github.com/thieuanhvan/t2dm-xai-bibliometric; Scopus CSV not redistributed due to provider terms of use but fully reproducible via documented query; PubMed records reproducible via `resubmission/05_run_all.py`) |

**Notes on Items Not Directly Applicable to Bibliometric Systematic Reviews**

**Item 11 (Risk of bias in individual studies).** Formal tools such as QUADAS-2 or CASP are designed to evaluate clinical-intervention or diagnostic-accuracy studies. The primary aim of the present review is to characterize the research-field structure through bibliometric and thematic analysis rather than to synthesize clinical evidence. Methodological characteristics of the 15 selectively reviewed high-citation papers (algorithm, XAI method, dataset, validation strategy) are transparently reported in Appendix A.

**Item 12 (Effect measures).** Clinical-effect measures (risk ratio, mean difference) are not applicable. The synthesis uses descriptive bibliometric measures: keyword-occurrence counts, publication-volume trends, citation counts, and ratio statistics (e.g., 53.3:1 XAI-to-KG disparity; 33.2% inter-database record overlap).

**Item 13d (Meta-analysis).** Meta-analysis was not performed because the included records are bibliographic metadata rather than clinical-outcome studies. The synthesis uses a rule-based thematic classifier (Section 3.3), which is transparent, reproducible, and consistent with bibliometric reporting norms.

**Item 15 / Item 22 (Certainty of evidence — GRADE).** GRADE is standard practice for systematic reviews of clinical interventions but is not directly applicable to bibliometric characterization of a research field. Internal-validity evidence is instead provided by the consistency of the principal finding (KG/GNN gap) across two independent databases with 33.2% record overlap, and by dual confirmation through quantitative bibliometric analysis and selective review of 15 highly cited papers.

**Item 24a (Prospective protocol registration).** The review was not prospectively registered in PROSPERO or OSF. PROSPERO currently restricts registration primarily to reviews with clinical-outcome synthesis, and the study was initially designed as a bibliometric analysis before being reframed as a PRISMA 2020-compliant systematic review in response to editorial guidance. The protocol is fully documented in Sections 3.1–3.4 and in the public code repository. Future related systematic reviews will be prospectively registered.
